# Systematic review investigating mHealth and digital health interventions for increasing vaccination uptake in 19 Sub-Saharan African countries: Recommendations for the malaria vaccine rollout

**DOI:** 10.1101/2025.04.24.25326272

**Authors:** Alex Bhattacharya, Chidera Mark-Uchendu, Christa Hansen, Jay Evans

**Affiliations:** University of Edinburgh, Usher Institute of Population Health Sciences and Informatics, Edinburgh, UK; Imperial College London, Institute of Global Health Innovation, London, UK; Imperial College London, Centre for Health Economics and Policy Innovation, London, UK; University College London, Centre for Teaching and Learning in Economics, London, UK

## Abstract

Mobile health and digital health (mHealth/DH) interventions have been shown to support immunisation programmes in Sub-Saharan Africa (SSA) and improve uptake of life-saving vaccines. As 19 SSA countries were targeted to begin rolling out the two new malaria vaccines (RTS,S/AS01 and R21/Matrix-M) in 2024, this systematic review aims to investigate which mHealth/DH interventions are most effective at increasing vaccination uptake (by assessing vaccination coverage and timeliness outcomes) in these 19 countries, and provide evidence-based recommendations on their use in the malaria vaccine rollouts.

As the malaria and DTP/Pentavalent vaccines share the same mode of delivery and vaccination schedule, mHealth/DH interventions targeting increase of DTP/Pentavalent vaccines were investigated as predictors for the malaria vaccines. Eight electronic databases were searched, along with several grey literature sources. A narrative synthesis was conducted with studies being grouped together by mHealth/DH intervention-type. Included studies were assessed for risk of bias using RoB2 and ROBINS-I, and certainty of evidence for each outcome was evaluated using the GRADE approach.

14 studies were included, comprising both randomised and non-randomised control trials. However, only 4 out of the 19 SSA countries were represented (Nigeria, Kenya, Burkina Faso and Cote D’Ivoire). All investigated interventions were immunisation appointment reminders. Generally, all mHealth/DH intervention-type subgroups were positively associated with vaccination coverage and timeliness. SMS-based interventions showed modest positive associations between intervention and outcomes, whereas voice-based phone call reminders, reported the strongest associations. The certainty of evidence ranged from very low to moderate depending on the intervention-type and outcome pairing. While the findings suggest that implementing mHealth/DH interventions, particularly those with voice-based components, in this context would likely improve vaccination coverage and timeliness, the limited certainty of evidence highlights the need for further high-quality research. Based on the presented evidence, a combined SMS and voice-based intervention is recommended for the malaria vaccine rollouts.

## Introduction

2024 was a monumental year for public health and the fight against malaria. The World Health Organization (WHO) has now approved two malaria vaccines for use (RTS,S/AS01 and R21/Matrix-M), and these are beginning to be rolled out in 19 countries (Benin, Burkina Faso, Burundi, Cameroon, Central African Republic, Chad, Cote d’Ivoire, Democratic Republic of Congo, Ghana, Guinea, Kenya, Liberia, Malawi, Mozambique, Niger, Nigeria, Sierra Leone, South Sudan and Uganda) across Sub-Saharan Africa (SSA) [1]. Therefore, understanding strategies to increase uptake of existing vaccines which share the same mechanism of delivery and three-dose schedules (such as the Diphtheria-Tetanus-Pertussis or Pentavalent (DTP/Pentavalent) vaccines) in this context, will provide valuable and potentially transferable evidence-based insights for the malaria vaccine rollout, and maximise the life-saving potential of these new vaccines [2]. There is increasing evidence to suggest that mobile health (mHealth) and digital health (mHealth/DH) interventions could optimise immunisation programmes and improve uptake of lifesaving vaccines [3,4], thus this systematic review will investigate mHealth/DH interventions for increasing vaccination uptake in the 19 SSA countries.

mHealth is an emerging field that involves the use of devices such as mobile phones and other wireless mobile devices to enhance medical and public health practices, and improve healthcare delivery, patient outcomes and health system efficiency [5,6]. mHealth interventions range from simple phone calls and short message service (SMS) messaging to more complex technologies like mobile applications, mobile data collection platforms and wireless data transmissions [7]. These support various activities, including facilitating health-related communications, remote disease surveillance, health education, data collection and analysis, healthcare worker (HCW) assistance, teleconsultations, research activities and streamlined patient management [6,8,9]. In contrast, digital health (DH) is a broad umbrella term encompassing eHealth (electronic health), which includes mHealth, in addition to areas such as ‘big data’ computing, genomics and artificial intelligence (AI) [10]. DH involves a wider range of technology, including electronic health records (EHR), telemedicine, online health education platforms and health information systems (such as the widely used District Health Information System2 (DHIS2)) [11]. These tools enable streamlined data collection, facilitating data-driven healthcare delivery decision-making [12].

As healthcare infrastructure is limited in certain SSA regions, mHealth/DH can support immunisation programmes where mobile phone penetration is high [13, 14]. Examples of immunisation-related mHealth interventions include SMS reminders to reduce missed appointments and ensure timely follow-ups, mobile data collection, and mHealth educational platforms to educate and address vaccine hesitancy [13]. Other DH tools are also useful in SSA immunisation programmes, including digital data management and analysis tools, tracking vaccination coverage, supply chain management, and telemedicine platforms for remote consultations or vaccinator training [4,15].

As the WHO targeted 19 SSA countries to begin the malaria vaccine rollouts in 2024, exploring strategies which will be effective at increasing uptake of comparable vaccines in this context is of public health significance [1]. Investigating strategies and interventions used in similar contexts is a key precursor in developing effective interventions [16]. Therefore, examining which mHealth/DH interventions have successfully increased vaccination uptake in comparable settings will allow evidence-based recommendations for the malaria vaccine rollout. Although disease-specific differences between malaria and DTP are acknowledged, this systematic review will offer valuable insights for intervention development.

Vaccination is a well-established tool for disease prevention and reducing mortality due to vaccine-preventable diseases (VPDs) [17]. Immunisations already prevent an estimated 3.5-5 million deaths annually from diseases such as diphtheria, tetanus, pertussis, and measles [17]. Both the RTS,S/AS01 and R21/Matrix-M malaria vaccines are administered to children at around 5months of age, through a 3dose primary schedule [18]. As the malaria vaccines and DTP/Pentavalent vaccine share the same mode of delivery (intramuscular injection) and immunisation schedule, this review will extrapolate outcomes of mHealth/DH interventions for increasing DTP/Pentavalent uptake, as predictors for what might be transferable to the malaria vaccine rollout. The DTP vaccine has already been used as a predictor for the malaria vaccines in economic evaluations of their cost-effectiveness [19]. Additionally using DTP/Pentavalent as a predictor for the malaria vaccines is appropriate as both require multiple doses. Vaccines requiring multi-doses for maximum protection require multiple appointments, therefore completing full immunisation schedules and achieving appropriate timeliness between doses is more challenging [20]. Non-adherence to schedules leads to dropouts which limits the vaccine’s protective benefits [20]. Therefore, exploring strategies for multi-dose schedule adherence will provide useful insights.

Malaria remains a leading cause of morbidity and mortality in Low-Middle-Income Countries (LMICs) [21]. Despite excellent progress being made in malaria control, malaria-attributed deaths remain unacceptably high [21]. 2022 saw an estimated 249million malaria cases globally and 608,000 deaths [21]. SSA accounted for 94% (233 million) of all cases and 95% (580,000) of malaria-attributed deaths (with 80% of deaths occurring in children under-5) [21]. The new malaria vaccines are a highly significant development for global vaccination efforts and offers opportunity to reduce malaria-attributed mortality [22]. The malaria immunisation programmes aim to reduce severe malaria incidence and mortality in infants and children under-5 exposed to high transmission rates [1]. The malaria vaccines have been predicted to be cost-effective (at 15-year time horizon) and have substantial public health impact by reducing clinical malaria cases and averting deaths [19, 23]. Given this predicted impact, finding strategies to optimise malaria vaccination uptake where the vaccines are introduced is a huge public health priority.

Four SSA countries accounted for almost half of all malaria-attributed deaths worldwide in 2022: Nigeria (26.8%), Democratic Republic of the Congo (12.3%), Uganda (5.1%) and Mozambique (4.2%) [21]. However, WHO announced 19 SSA countries will be rolling out the malaria vaccines in 2024, and these will be this review’s focus [1, 24]. Investigating strategies to increase vaccination uptake in this specific context is crucial for maximising the life-saving potential of the vaccines.

Challenges facing SSA immunisation programmes are well documented and relate to socio-economic, infrastructural and political factors [25]. Examples include: inadequate healthcare infrastructure presenting logistical challenges which disrupt vaccine cold chain storage and their distribution and uptake [26], supply chain issues which can result in vaccine wastage or ‘stockouts’ resulting in unavailability of required vaccines [27, 28], and vaccine hesitancy due to misinformation or cultural beliefs [29]. Economic constraints are also a key barrier, with many SSA countries relying on external funding to execute national immunisation programmes [25]. On an individual level, poor adherence to immunisation appointments means essential immunisation schedules are not completed, reducing efficacy and life-saving benefits of the vaccines [30]. Although some of these challenges are beyond the scope of mHealth/DH interventions, these technologies can offer solutions for poor adherence in appointments vaccine hesitancy and enhancing supply chain management [13, 31, 32].

Although mobile phone affordability is a barrier for many in SSA, huge projected growth in SSA mobile phone use over the coming decades presents a significant opportunity for mHealth/DH where healthcare infrastructure may be lacking. Despite this, equity considerations relating to the ‘digital divide’ are essential during intervention development [33].

Initial scoping searches of mHealth/DH interventions for increasing vaccination uptake identified four categories for immunisation programme optimisation. These are: communication technology (such as SMS appointment reminders), stock management mobile applications, surveillance or data analysis tools and electronic immunisation registries (EIR) [13, 34, 35, 36]. Leveraging these technologies could be hugely advantageous for improving immunisation programmes and related health outcomes. Most published experimental literature related to mHealth/DH and communication technology focusing on appointment scheduling and reminders.

Several systematic reviews on similar topics were discovered [13, 34, 37]. However, this review will seek to answer a different research question, search for both mHealth/DH interventions and is explored from a novel perspective of what might be effective for the malaria vaccine rollout. Additionally, given the frequent introduction of new mHealth/DH technologies and research, regular reviews are justified to update the evidence base [38]. The results will offer future generalisability and recommendations on which interventions could be most effective for the malaria vaccine rollout.

The review will seek to answer the research question: ‘Which mHealth or digital health interventions have proved most effective in increasing childhood vaccination uptake (of DTP/Pentavalent vaccine) in the 19 SSA countries rolling out malaria vaccine in 2024?’, by addressing three outlined objectives: **(1)** Identify current up-to-date (as of January 2025) evidence of mHealth/DH interventions for increasing vaccination uptake and coverage of DTP/Pentavalent vaccines (as a predictor for the malaria vaccines) in the outlined 19 SSA countries, **(2)** Report on any other factors from included studies relating to uptake of childhood immunisations, for example timeliness of vaccination, missed opportunity for vaccination or vaccine wastage in the selected SSA countries, and **(3)** Provide strategic evidence-based recommendations based on review’s findings on how mHealth/DH interventions can be used to improve vaccination uptake in the malaria vaccine rollout context. Other implementation challenges, risks or mitigating factors will also be considered here.

## Methods

This systematic review of quantitative articles was guided by the Cochrane Methodology and followed the PRISMA reporting guidelines [39, 40]. The completed PRISMA checklist is included in **Supplementary Information** (**S1**). Initial scoping searches to assess volume of literature on the topic were conducted on Google Scholar and PubMed. The review was conducted in line with a predefined protocol which was registered on PROSPERO in October 2024 (CRD42024587428).

The PICOS (population, intervention, comparison, outcome and study design) framework was used to structure several review stages including the search strategy, inclusion/exclusion criteria, study selection process and data extraction [41]. We adapted this framework to include an additional ‘S’ representing setting given the review’s specific geographic context. Therefore, our PICOSS framework used is shown in **Supplementary Information** (**S2**). The outcomes of interest were selected to provide a comprehensive understanding of mHealth/DH interventions effect on vaccination uptake. The review investigated both key parameters relating to vaccination uptake: coverage (indicating the proportion of study population under investigation that received DTP/Pentavalent vaccine), and timeliness (indicating whether vaccines were administered at recommended times ensuring optimal effectiveness) [42].

Eight electronic databases were selected based on their relevance for global health research. These were MEDLINE (Ovid), Global Health (Ovid), Embase (Ovid), Scopus, Web of Science, Cochrane CENTRAL, WHO’s African Index Medicus (AIM) and African Journals Online (AJOL). The search strategy development was done in collaboration with University of Edinburgh’s College of Medicine and Veterinary Medicine’s Academic Support Librarian. The search strategy for each database is shown in **Supplementary Information** (**S3**). Final searches were conducted on January 17th, 2025. Additionally, a small grey literature search was conducted to reduce potential publication bias, the sources searched were OpenHIA [43], OpenMRS [44], and WHO’s mHealth/DH working group publications [45]. Finally, to ensure no key studies were missed, all the included studies of identified similar reviews were assessed for eligibility if not already included.

All retrieved studies were imported into Covidence review management software which was used to facilitate the review process. A PICOSS-based inclusion and exclusion criteria guided the screening process (**S2**). AB and CMU conducted title and abstract, and full-text screening, independently, with JE resolving any arising conflicts. Study selection was presented using an adapted PRIMSA flowchart [40]. As experimental epidemiological studies (RCTs and non-RCTs) are the gold standard for testing intervention effectiveness, only these study types were included [46]. Data extraction was also conducted by two researchers independently (AB and CMU). AB and CMU also conducted an independent risk of bias assessment of all included studies to assess each included study’s methodological rigour. Cochrane’s Risk of Bias2 (RoB2) tool was used for assessing RCTs [47], whilst Cochrane’s Risk of Bias in Non-randomised Studies (ROBINS-I) tool was used to assess non-RCTs [48]. Additionally, a discrete certainty of evidence assessment was conducted to assess confidence in the review’s findings for each outcome (mHealth/DH intervention effect on: (1) vaccination coverage and (2) vaccination timeliness) using the GRADE (Grading of Recommendations Assessment, Development and Evaluation) framework [49, 50].

### Data analysis and narrative synthesis

As this paper aims to identify which interventions are most effective at increasing vaccination uptake in the specific context, the analysis aimed to evaluate the relative effectiveness of each included mHealth/DH intervention on vaccination coverage and timeliness. AB and CH conducted an assessment of the extracted data and made decision on synthesis approach. Due to extremely high heterogeneity reported (**Figure 3** and **4**), a narrative synthesis guided by synthesis without meta-analysis (SWiM) reporting guidelines was conducted [51]. This approach was further supported by very high heterogeneity reported in similar meta-analysis studies [34, 37]. For the narrative synthesis, studies were grouped together by mHealth/DH intervention-type. These intervention-type groups were: ‘SMS-Only’, ‘SMS-Plus’. ‘SMS and/or Voice Messages or Phone calls’, ‘Phone calls only’, and ‘Electronic Immunisation Alert Wristband’. A description of each group and the studies included in each is shown in **Supplementary Information** (**S4**). The synthesis method entailed a description of positive or negative associations, a consideration of point estimate precision, and each study’s risk of bias. Additionally, for narrative synthesis best practice suggests that syntheses are accompanied by discrete certainty of evidence assessment [49-51]. As mentioned, the certainty of the evidence for each intervention-outcome pairing using the GRADE framework was conducted. This considered five domains: risk of bias, inconsistency, indirectness, imprecision, and publication bias. AB and CMU independently conducted the assessments, resolving any discrepancies through discussion. Final certainty ratings were classified as high, moderate, low, or very low. Due to a lack of variation in results due to study design, it was deemed appropriate to analyse findings from both RCT and non-RCT study designs together.

Forest plots were created using Jamovi statistical software to provide visual representation of the review’s quantitative findings and to support the narrative synthesis. These only presented data on the third DTP/Pentavalent dose (DTP/Pentavalent3). Using the final dose provided a robust indicator of mHealth/DH intervention effectiveness on vaccine coverage and timeliness, and indicated whether the vaccination schedule was completed, thus allowing assessment of intervention effectiveness on schedule adherence. Each included intervention was analysed individually, therefore multi-arm studies provided multiple point estimates. This allowed a visual comparison of all included interventions. It should be noted that each intervention from the multi-arm RCTs or non-RCTs was compared against the same control, however, this does not bias the results as pooled estimates were not calculated.

The standardised synthesis metric was logarithm of the odds ratio (logOR), which provided information on associations between mHealth/DH intervention and outcome (vaccination coverage or timeliness). All values were transformed to LogOR, as this metric provided a more robust visualisation of strength and certainty of reported associations [52, 53]. LogORs were calculated from the data extracted from included studies, the raw data used to calculate point estimates is shown in **Supplementary Information** (**S6**). 95% confidence intervals were provided for each point estimate, along with heterogeneity statistics (including I^2^).

## Results

The PRISMA flow chart outlines the study selection process (**Figure 1**). The full search identified 6424 records, however, following full-text screening only 14 studies met the inclusion criteria and were included in the study [54-67]. The full list of included studies is shown in **Supplementary Information** (**S4**).

**Figure 1.**
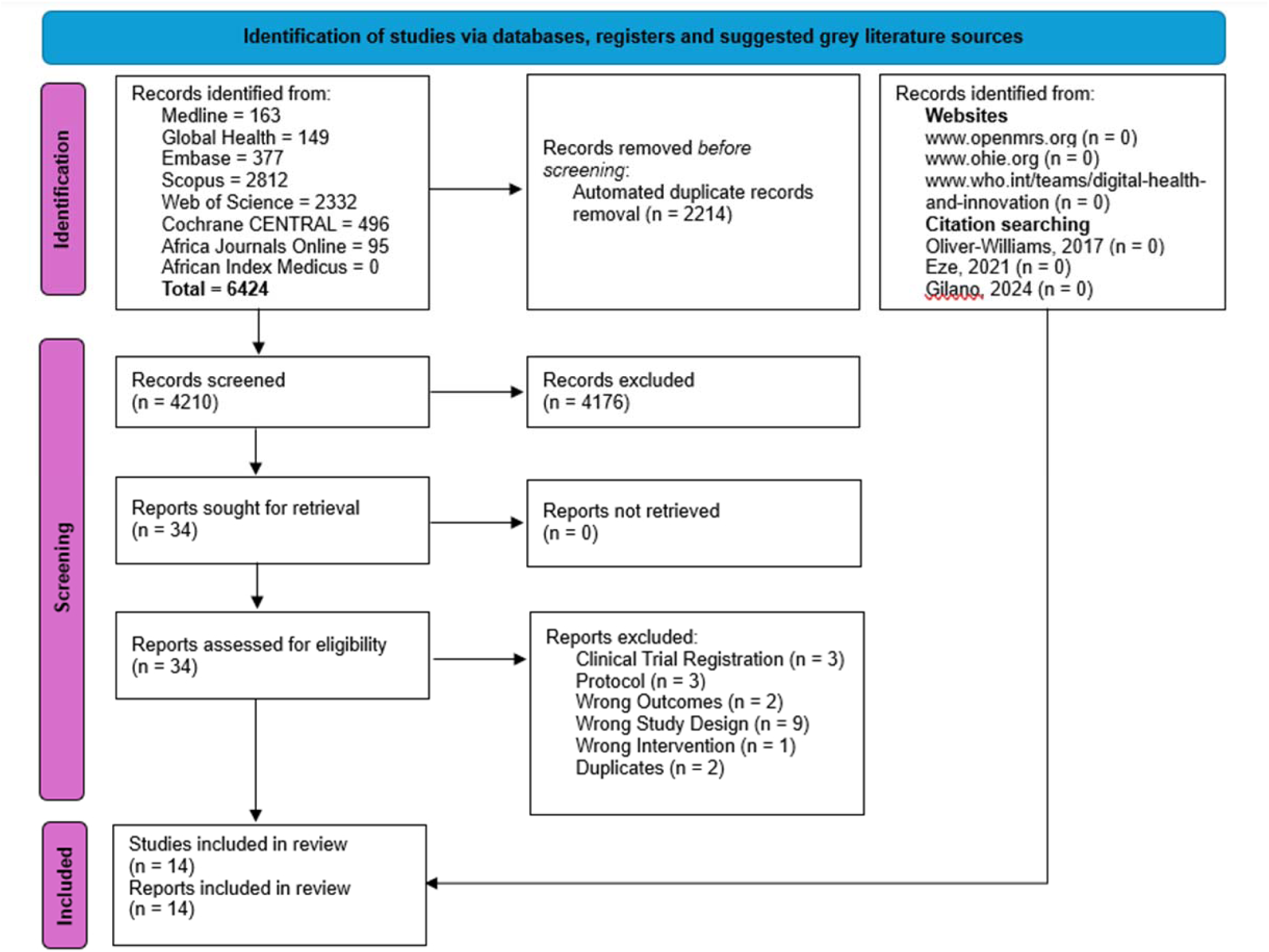
PRISMA flow chart outlining the study selection process for this systematic review. Adapted from Page *et al* (2021) [40].

### Study Characteristics

**Table 1** summarises the characteristics of the 14 included studies [54-67]. 10 of the included studies were RCTs [54-60, 62, 65, 67], with the remaining 4 studies comprising non-RCTs [61, 63, 64, 66]. The majority of studies were conducted in West Africa (12 studies) [54-58, 61-67], and only 4 of the 19 SSA countries targeted for malaria vaccine rollout (Nigeria [54, 55, 57, 58, 61-64, 66, 67], Burkina Faso [65], Cote D’Ivoire [56] and Kenya [59, 60]) were represented in the study. All the interventions included related to appointment reminders to encourage and remind mothers/caregivers to bring their children to immunisation clinics to receive DTP/Pentavalent doses. As each intervention was analysed independently, the number of mHealth/DH interventions and the type of interventions investigated in this review is shown in **Figure 2**.

**Figure 2.**
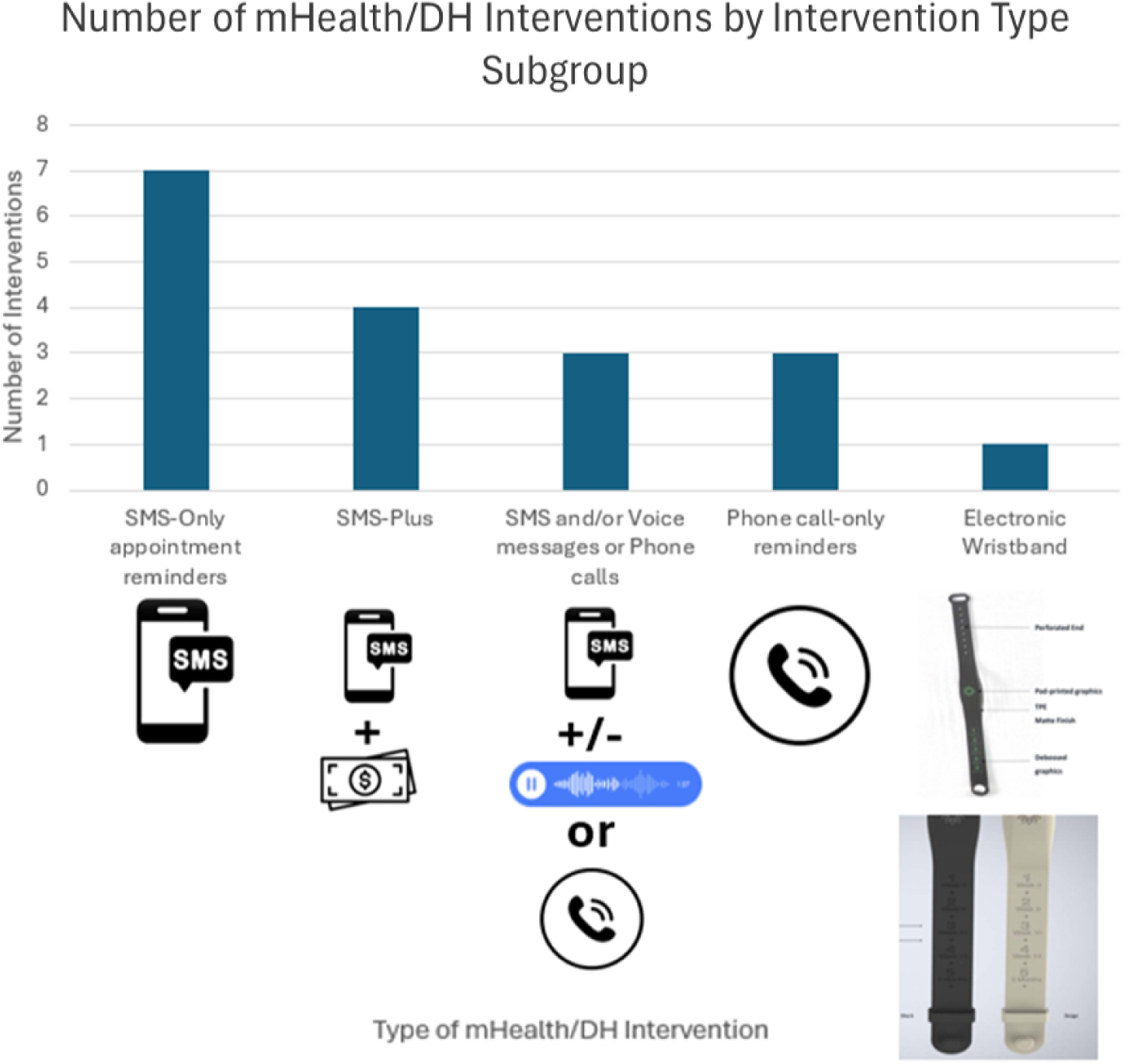
The number of interventions investigated in this review by intervention-type subgroup and pictograms of each intervention. Note that individual interventions in multiple-arm RCT and non-RCTs have been investigated separately. (Created on Microsoft Excel and PowerPoint, pictures taken from canva.com and Sampson *et al* (2023) [64]).

**Table 1.** Study Characteristics of the 14 included studies.

| Author and Year | Country (Region) | Urban or Rural Setting | Study Design | mHealth/DH Intervention | Intervention Details | Control Details | Study Population | Follow Up Length | Outcomes Measured (Coverage or Timeliness) | DTP or Pentavalent Vaccine | Additional Notes |
| --- | --- | --- | --- | --- | --- | --- | --- | --- | --- | --- | --- |
| Brown and Oluwatosin, 2017 [54]. | Nigeria (West Africa) | Urban | RCT (2-arm) (Clustered design) | 'Phone call-only' appointment reminders | Intervention group received 1x phone call reminder 2days before their child's next immunisation appointment. If appointment was missed, then 'recall' phone call was made. | Usual care - no phone call reminders | 595 mother/infant pairs (295 in the intervention group and 300 in the control group). Children were aged 0-12weeks at recruitment. | 13 months | Immunisation compliance rate (Considers the required number of doses at appropriate age and recommended intervals [schedule adherence/timeliness]) (%). **Categorised as coverage.** | DTP vaccine | Used same study population as Brown *et al* (2016). Group **A+C** in Brown *et al* (2016) = Intervention group. Group **B+D** in Brown *et al* (2016) = Control. RCT registered and results analysed retrospectively. |
| Brown *et al*, 2016 [55]. | Nigeria (West Africa) | Urban | RCT (4-arm) (Clustered Design) | 'Phone call-only' appointment reminders | **Group A** received phone calls 2days before and 1day before the child's immunisation appointment. 'Recall' phone calls were made if an appointment was missed. **Group B** no mHealth/DH intervention so not included. **Group C** received both phone call reminders and vaccinator/HCW training on immunisation theory. | **Group D** received usual care - no intervention | 595 mother/infant pairs total study population (295 in the intervention groups [148 in **A** and 147 in **C**] and 150 in the control group [**D**]. **Group B** not included. Children were aged 0-12weeks at recruitment. | 12 months | Immunisation schedule completed or not completed (All essential immunisations including 3x doses of DTP/Pentavalent) (%; RR and OR). **Categorised as coverage.** | DTP vaccine | This report is the larger study mentioned above in Brown and Oluwatosin (2017) |
| Dissieka *et al*, 2019 [56]. | Cote D'Ivoire (West Africa) | Both | RCT (2-arm) (Individual) | Automated SMS or Voice message appointment reminders | Intervention group received either SMS or voice message prior to each scheduled immunisation visit. 2x additional reminders were sent if appointment was missed. Reminders were automatically sent out. Mothers could choose whether they received SMS or phone call, and language of reminders. | Usual care - no intervention | 1596 mother/infant pairs (798 in intervention group and 798 in the control group). 736 were from Urban setting, 560 were from Semi-urban setting, 300 were from Rural setting). | 12 months | Attendance at each of the 3x Pentavalent vaccination appointment (% and aOR). **Categorised as coverage.** | Pentavalent vaccine |  |
| Ekhaguere *et al*, 2019 [57]. | Nigeria (West Africa) | Semi-rural | RCT (2-arm) (Individual ) | Automated SMS and Voice message appointment reminders | Intervention group received automated phone call and text messaging appointment reminders, sent out 2days, and 1day, before the scheduled appointment for 3x Pentavalent doses. Text messages were in English, whereas phone calls were in English and Yoruba. | Usual care - no intervention. Child immunisation card listed when child was to receive immunisations . | 600 mother/infant pairs (300 in the intervention group and 300 in the control group). | 12 months | Proportion of infants that received Pentavalent dose1, dose 2 and dose 3 (% and RR). **Categorised as coverage.** | Pentavalent vaccine |  |
| Eze and Adeleye, 2015 [58]. | Nigeria (West Africa) | Urban | RCT (2-arm) (Individual ) | 'SMS-only' appointment reminders | Intervention group was sent SMS appointment reminder messages 1day before the child's next immunisation appointment. 'Recall' SMS messages were sent 1day before the next immunisation session when an appointment was missed. | Usual care - no intervention | 1001 mother/infant pairs (500 in intervention group and 501 in control group, 96 were lost to follow up so 905 were included in analysis (452 in intervention group and 453 in control group). | 18 weeks | DTP3 Coverage (%) and Timeliness of receipt (%). **Both coverage and timeliness.** | DTP vaccine |  |
| Gibson *et al*, 2017 [59]. | Kenya (East Africa) | Rural | RCT (4-arm) (Clustered design) | Automated SMS reminders and SMS plus cash incentives ('SMS-Plus') | **Group A** received only SMS appointment reminders.<br>**Group B** received SMS appointment reminders + cash incentive of 75KES, **Group C** received SMS appointment reminders + cash incentive of 200KES. All reminders were sent 1day before scheduled immunisation appointment (6weeks, 10weeks, 14weeks for 3 pentavalent doses). | **Group D.** Usual care - no SMS reminders or financial incentives. | 2018 mother/infant pairs (489 in the control group, 476 in SMS only group, 562 SMS+75KES group and 491 SMS+200KES group). | 12 months | Proportion of fully immunised children at 12months (included 3x Pentavalent doses plus BCG, 3x OPV and MCV) (% and RR) and timeliness of receipt (within 2weeks of recommended schedule) (% and RR). **Both coverage and timeliness.** | Pentavalent vaccine |  |
| Haji *et al*, 2016 [60]. | Kenya (East Africa) | Both | RCT (3-arm) (Clustered design) | Automated 'SMS-only' appointment reminders | Intervention group received 2x SMS appointment reminders 2days before child's next appointment for 2nd and 3rd DTP doses. The 1st message reminded caretaker of the next due date of appointment and where to attend. The second message reminded caretakers that appointment was that day. Messages were in both Kiswahili and English. | Usual care - non-SMS. Appointment was recorded in child's health book. | 1116 mother/infant pairs (372 in SMS intervention group, 372 in Sticker reminder group [Non-mHealth/DH intervention so not included] and 372 in control group). | 14 weeks | Penta2 and Penta3 coverage (%), and dropout rate (%). **Categorised as coverage.** | Pentavalent vaccine | Another non-mHealth/DH intervention was included in this study of 'sticker reminders', however, was not included in this study. |
| Ibraheem *et al*, 2021 [61]. | Nigeria (West Africa) | Urban | Non-RCT (4-arm) (Clustered design) | Phone call appointment reminders (**A**), 'SMS-only' appointment reminders (**B**), or SMS health education ('SMS-Plus') (**C**) | **Group A** received phone call appointment reminders. **Group B** received SMS text message appointment reminders. **Group C** received health education SMS messages. **Group A** and **B** received reminders day before scheduled appointment. Phone call reminders were in English and/or Yoruba. SMS health education messages were sent at week5, week9 and 8months. | **Group D** received usual care - no intervention | 560 mother/infant pairs (140 in **Group A** [phone call reminder group], 140 in **Group B** [SMS reminder intervention group], 140 in **Group C** [SMS health education intervention group], 140 in **Group D** [control group]). | 9 months | Vaccination coverage (%), proportion of infants that received timely Penta1, Penta2 and Penta3 (%) and likelihood of presentation for appointment (OR). **Both coverage and timeliness.** | Pentavalent vaccine | All intervention subgroups represented by this study |
| Kawakatsu *et al*, 2020 [62]. | Nigeria (West Africa) | Urban | RCT (2-arm) (Individual) | Automated 'SMS-only' appointment reminders | Intervention group received SMS appointment reminders in English, 2days before an upcoming appointment. In the event of a no show, an additional SMS reminder was sent 7days after the original appointment date. | Usual care - no intervention | 8337 mother/infant pairs attending appointments at 31 primary healthcare centres, 4893 were in intervention group and 3444 were in control group. | 3 months | Return rate for subsequent immunisation appointment (% and aOR). **Categorised as timeliness.** | Pentavalent vaccine |  |
| Oladepo *et al*, 2020 [63]. | Nigeria (West Africa) | Rural | Non-RCT (2-arm) (Clustered design) | Automated SMS immunisation education/reminders ('SMS-Plus') | SMS immunisation education messages sent 3x/week for 10months. Messages focused on importance of keeping to immunisation appointment, benefits of keeping routine appointments, benefits of timely and full completion of all basic essential immunisations and consequences of refusal/non-completion. Messages were in English but also translated in Yoruba, Ijaw, Hausa and Igbo. | Usual care - no intervention. However, were given flyers on the importance of adequate child nutrition and growth monitoring. | 3500 mother/infant pairs (1750 in intervention group and 1750 in control group). | 10 months | Completion of all immunisations (%) and coverage of Penta1, Penta2 and Penta3 (%). **Categorised as coverage.** | Pentavalent vaccine |  |
| Sampson *et al*, 2023 [64]. | Nigeria (West Africa) | Both | Non-RCT (2-arm) (Clustered design) | Wearable electronic immunisation appointment alert wristband | The electronic immunisation alert wristband was worn by caregiver and programmed to flash a red light in the lead up to the child's next immunisation appointment and to the day of appointment. The device had undergone successful pilot testing. | Usual care - no intervention | 757 caregivers were in the intervention group, 190 were in the control group. | 9 months | Timeliness of immunisation rate (%) and reduction in dropout rate (%). **Categorised as timeliness.** | Pentavalent vaccine |  |
| Schlumberger *et al*, 2015 [65]. | Burkina Faso (West Africa) | Urban | RCT (2-arm) (Individual) | Automated 'SMS-only' appointment reminders | SMS appointment reminders sent before each of the eight immunisation sessions. Messages communicated that the caregiver was expected at the location of the appointment. | Usual care - no intervention | 523 Mother/infant pairs (253 in the intervention group and 268 in control group). | 5 months | Penta1, Penta2 and Penta3 coverage rate (%) and immunisation timeliness rate (%). **Both coverage and timeliness.** | Pentavalent vaccine |  |
| Yunusa *et al*, 2022 [66]. | Nigeria (West Africa) | Urban | Non-RCT (2-arm) (Clustered design) | 'SMS-only' appointment reminders | Intervention group received SMS reminders 3days before their child's next immunisation appointment, and on the day of appointment. Therefore, each mother/caregiver received 2x SMS for each of 3x Pentavalent doses. | Usual care - no intervention | 541 Mother/infant pairs (271 in the intervention group and 270 in control group). | 6 months | Penta1, Penta2 and Penta3 coverage rate (% and OR). **Categorised as coverage.** | Pentavalent vaccine |  |
| Yunusa *et al*, 2024 [67]. | Nigeria (West Africa) | Urban | RCT (2-arm) (Clustered design) | SMS and phone call reminders | Intervention group was sent 6x SMS and 3x phone call reminders at specific time intervals during study timeframe. 1st SMS was welcoming message, 2nd & 3rd were sent to participants in 4th & 5th week with their child's appointment dates. 4th, 5th & 6th SMS reminders were sent 3x days before the scheduled date that their baby was due for immunisation (6, 10 & 14 weeks after birth). | Usual care - no intervention | 554 Mother/infant pairs (277 in intervention group and 277 in control group, 18 were lost to follow up), so 536 Mother/infant pairs were analysed (275 in intervention group and 261 in control group). | 14 weeks | Penta1, Penta 2 and Penta3 coverage (%) and Timeliness of immunisation (%). Both coverage and timeliness. | Pentavalent vaccine | Article in press |

Note Penta1, Penta2 and Penta3 refers to 1st, 2nd and 3rd Pentavalent dose respectively. RR = Risk Ratio, OR = Odds Ratio, aOR = adjusted Odds Ratio, KES = Kenyan Shillings, BCG = Bacillus Calmette-Guérin vaccine, DTP = Diphtheria, Tetanus, and Pertussis vaccine, MCV = measles-containing vaccine, OPV = Oral Polio vaccine.

### Outcomes under investigation

The two key outcomes investigated relating to vaccination uptake are vaccination coverage and timely receipt of doses. This review investigated mHealth/DH intervention effect on both these different but complementary outcomes to provide a complete understanding of vaccination uptake. Including mHealth/DH intervention effective on coverage helps to identify the intervention effectiveness on reaching the target population [42]. Whereas including intervention effect on timeliness is important for understanding how effective interventions are for ensuring vaccines are administered within correct time window, allowing for optimal immunity [42]. 7 studies included outcomes categorised as only relating to vaccination coverage [54-57, 60, 63, 66]. 5 studies measured both coverage and timeliness [58, 59, 61, 65, 67]. While only two studies only reported outcomes relating to timeliness [62, 64].

### Study Findings

**Table 2** shows the key findings from the included studies. The two key parameters (vaccination coverage and timeliness), align with **Study Objectives 1** and **2**. As mentioned each intervention from the 14 studies has been presented individually, therefore were 17 interventions relating to coverage and 12 interventions relating to timeliness are presented.

**Table 2.** The key findings from the 14 included studies relating to the 3 study objectives.

| Author and year | Outcomes Measured (Coverage or Timeliness) | Results Relating to Study Objective 1 (Vaccination Coverage) | Results relating to Study Objective 2 (Vaccination Timeliness) | Additional Results relating to Study Objective 3 | LogOR presented in Forest plots (Only 3<sup>rd</sup> DTP/Pentav alent dose) | Overall Conclusions |
| --- | --- | --- | --- | --- | --- | --- |
| Brown and Oluwatosi, 2017 [54]. | Immunisation compliance rate (Considers the required number of doses at appropriate age and recommended intervals) (%). **Categorised as coverage.** | Immunisation compliance rate. **Intervention Group:** 79.2% **Control Group:** 46.4% (p < 0.001*). | N/A | 98.2% of mothers/caregivers (n=584) in the study population agreed to receive the immunisation reminder/recall phone calls. Therefore, was an acceptable intervention. | **Coverage:** LogOR 1.49 (95% CI 1.13-1.85)<br>**Timeliness:** N/A | The results indicate that phone call reminders by health workers to mothers/caregivers can enhance immunisation compliance to the 3x dose schedule of DTP. * = statistical significance difference between the groups (p<0.05). |
| Brown *et al*, 2016 [55]. | Immunisation schedule completed or not completed (All essential immunisations including 3x doses of DTP/Pentavalent) (%), RR and OR). **Categorised as coverage.** | Immunisation schedule completion rate: **Group A (Phone call reminders)** = 98.6%. **Group C (Phone call reminders + Vaccinator training)** = 97.3%. **Group D (Control)** = 57.3%. RRs: **Group A vs Group D** = 1.72 (95% CI 1.5-1.98), **Group C vs Group D** = 1.70 (95% CI 1.47-1.95). ORs: **Group A** = 54.33 (95% CI 13.68-464.56, p<0.0001*), **Group C** = 26.6 (95% CI 9.32-103.13, p<0.0001). | N/A | Vaccinator training alone (**Group B**) only marginally different to control group (RR 1.22, 95% CI 1.03-1.45). Group B compared to D, OR was 1.58 (95% CI 0.96-2.59) was not significant. | **Coverage:** (A) LogOR 3.99 (95% CI 2.56-5.43) (C) LogOR 3.28 (95% CI 2.24-4.33)<br>**Timeliness:** N/A | The results show phone call reminders are associated with highest completion rate of the essential childhood immunisation schedule. Combining the intervention with training of the vaccinators was not superior to only using phone call reminders. This supports the implementation of phone call reminders in immunisation programmes to improve immunisation completion rates. * = statistical significance (p<0.05). |
| Dissieka *et al*, 2019 [56]. | Attendance at Penta1, Penta2 and Penta3 dose appointments (% and aOR). **Categorised as coverage.** | Immunisation appointment attendances. **Penta1:** Intervention group = 691 (86.6%), Control group = 607 (76.1%), (aOR = 2.85 (95% CI 1.85-4.37) (p< 0.001*). **Penta2:** Intervention = 646 (81%), Control = 537 (67.3%), aOR = 2.8 (95% CI 1.88-4.17) (p< 0.001*). **Penta3:** Intervention = 592 (74.2%), Control = 465 (58.3%), aOR = 2.68 (95% CI 1.84-3.91) (p< 0.001*). | N/A | In intervention Group 84.6% (n=675) chose to receive a voice message reminder and 15.4% (n=123) chose to receive SMS message reminders prior. Voice messages were preferred by 97.7% (n=293) of mothers in rural areas. No significant difference between whether SMS or Voice message was used. | **Coverage:** LogOR 0.72 (95% CI 0.51-0.93)<br>**Timeliness:** N/A | Results show that sending voice call or SMS appointment reminders increase attendance at each immunisation appointment. Majority of mothers chose voice messages in rural areas, suggesting that providing an option of SMS or voice call reminders would be most useful (particularly in more rural areas where literacy rates may be lower). * = statistical significance (p<0.05). |
| Ekhuaguer *et al*, 2019 [57]. | Proportion of infants that received Penta1, Penta2 and Penta3 doses (% and RR). **Categorised as coverage.** | Pentavalent vaccine coverage. **Penta1:** Intervention = 285 (95%), Control = 289 (97%), RR was 0.98 (95% CI 0.95-1.02) p=0.31. **Penta2:** Intervention = 276 (92%), Control = 278 (93%), RR was 0.99 (0.95-1.04) p=0.76. **Penta3:** Intervention = 257 (86%), Control = 244 (81%), RR 1.05 (0.98-1.13) p=0.15. | Pentavalent timeliness (receiving each within 1 week of recommended time). **Penta1:** Intervention = 257 (86%), Control = 250 (83%), RR was 1.03 (95% CI 0.96-1.10, p=0.43). **Penta2:** Intervention = 256 (85%), Control = 252 (84%), RR was 1.02 (0.95 - 1.09) p=0.65. **Penta3:** Intervention = 253 (84%), Control = 233 (78%), RR was 1.09 (1.01-1.17, p=0.04*). | Completing all immunisations by 12months of age was also recorded. Intervention = 171 (57%), Control = 140 (47%), RR was 1.22 (1.04-1.43, p=0.01*). | **Coverage:** LogOR 0.32 (95% CI - 0.12-0.75)<br>**Timeliness:** LogOR 0.46 (95% CI 0.05-0.88) | No statistically significant difference was found between intervention group and control group for coverage of the 3x doses of Pentavalent vaccine (only significant difference in coverage was registered for MCV but this is not included in study). However, paired automated calls with SMS reminders significantly improved the proportion of infants that completed all routine immunisations by 12 months and significantly improved the timeliness of Penta3. * = statistical significance (p<0.05). |
| Eze and Adeleye, 2015 [58]. | DTP3 Coverage (%) and Timeliness of receipt (%). **Both coverage and timeliness.** | **DTP3** coverage = 69% for intervention group, 60.3% in control group. | DTP3 timeliness of receipt. Children were categorised into 'early' or 'delayed' (DTP3 is scheduled for 14th week, early = received DTP before 18weeks, missing appointment before 18weeks was categorised as delayed).<br>**Early DTP3:** Intervention = 312 (69%), Control = vs 273 (60.3%), OR = 1.468 (95% CI 1.103-1.955). | N/A | **Coverage:**<br>LogOR 0.38 (95% CI 0.11-0.66)<br>**Timeliness:**<br>LogOR 0.38 (95% CI 0.11-0.66) | Automated SMS and Voice message appointment reminders did increase the coverage of DTP3 vaccine; however, it is unclear whether this difference was statistically significant. The timeliness results showed that those in the intervention group were more likely to be 'early' than 'delayed' meaning the intervention did improve vaccination timeliness. |
| Gibson *et al*, 2017 [59]. | Proportion of fully immunised children at 12months (included 3x Pentavalent doses plus 1x BCG, 3x OPV doses and 1x MCV) (%) and RR) and timeliness of receipt (within 2weeks of recommended schedule) (%) and RR). **Both coverage and timeliness.** | Full immunisation at 12months.<br>**Group A** ('SMS-only') = 332 (86%), RR 1.04 (95% CI 0.97-1.12, p=0.29). **Group B** (SMS+75KES) = 383 (86%), RR 1.04 (95% CI 0.96-1.11, p=0.33). **Group C** (SMS+200KES) = 364 (90%), RR was 1.09 (95% CI 1.02-1.16, p=0.0014*). **Group D** (Control) = 296 (82%). Pentavalent dose coverage.<br>**Penta1:** All groups achieved 100%.<br>**Penta2:** **Group A** = 383 (99%), RR 1.00 (95% CI 0.98-1.101, p=0.77). **Group B** = 442 (99%), RR 1.00 (95% CI 0.99-1.02, p=0.85). **Group C** = 404 (99.5%), RR 1.01 (95% CI 0.99-1.02, p=0.42). **Group D** = 356 (99%).<br>**Penta3:** **Group A** = 375 (97%), RR 0.98 (95% CI 0.96-1.01, p=0.20). **Group B** = 439 (98%), RR 1.00 (95% CI 0.98-1.02, p=0.82). **Group C** = 401 (99%), RR 1.00 (0.99-1.02, p=0.58). **Group D** = 353 (98%). | Timeliness of Pentavalent doses (within 2weeks of recommended schedule).<br>**Penta1:** **Group A** = 347 (89%), RR 0.98 (95% CI 0.94-1.03, p=0.47). **Group B** = 412 (92%), RR 1.01 (95% CI 0.97-1.06, p=0.55). **Group C** = 377 (93%), RR 1.02 (95% CI 0.98-1.06, p=0.40). **Group D** = 328 (91%).<br>**Penta2:** **Group A** = 320 (82%), RR 0.98 (95% CI 0.92-1.05, p=0.54). **Group B** = 387 (87%), RR 1.03 (0.97-1.09, p=0.31), **Group C** = 359 (88%), RR 1.05 (95% CI 0.99-1.11, p=0.093). **Group D** = 303 (84%).<br>**Penta3:** **Group A** = 288 (74%), RR 1.01 (95% CI 0.91-1.11, p=0.90). **Group B** = 354 (79%), RR 1.07 (95% CI 0.98-1.17, p=0.16). **Group C** = 337 (83%), RR 1.12 (95% CI 1.03-1.22, p=0.0092*). **Group D** = 367 (74%). | Adding financial incentives did show significant improvement in timeliness of MCV (however not reported in this study). | **Coverage:**<br>**(A)** LogOR -0.56 (95% CI -1.49-0.37)<br>**(B)** LogOR 0.22 (95% CI -0.84-1.28)<br>**(C)** LogOR 0.46 (95%CI -0.69-1.62)<br>**Timeliness:**<br>**(A)** LogOR 0.00 (95% CI -0.32-0.33)<br>**(B)** LogOR 0.29 (95% CI -0.04-0.62)<br>**(C)** LogOR 0.53 (95%CI 0.18-0.88) | The results show that combining SMS reminders plus cash incentives ('SMS Plus' intervention) were modestly effective at improving the proportion of children fully vaccinated by 12 months of age and SMS reminders (only incentivising with 200KES offered statistical significance). No significant difference was reported between the groups for Penta1, Penta2 or Penta3 coverage. These interventions were more effective at improving timeliness of Penta doses, with timeliness of receipt of Penta3 in Group C (SMS+200KES) showing a significant difference, indicating larger cash amount is more favourable for this outcome. * = statistical significance (p<0.05). |
| Haji *et al*, 2016 [60]. | Penta2 and Penta3 coverage (%), and dropout rate (%). **Categorised as coverage.** | Pentavalent dose coverage.<br>**Penta2:** Intervention= 365 (98%), Control = 340 (91%).<br>**Penta3:** Intervention = 359 (96%), Control = 309 (83%). | Dropout Rate.<br>**Intervention:** 365 (98%) received Penta2 and 359 (96%) received Penta3 dose (p = 0.4).<br>**Control:** 340 (91%) received Penta2 and 309 (83%) received Penta3 (p<0.001*). There was a significant increase in dropouts in control between Penta 2 and 3 but not in intervention. | N/A | **Coverage:**<br>LogOR 1.73 (95% CI 1.11-2.34)<br>**Timeliness:**<br>N/A | The results show SMS reminders were effective in reducing dropouts for vaccinations in the selected Kenyan districts. The vaccination coverage was higher among those receiving SMS reminder than those receiving routine reminders. * = statistical significance (p<0.05). |
| Ibraheem *et al*, 2021 [61]. | Penta1, Penta2, Penta3 coverage and timeliness (%). Penta1, Penta2 and Penta3 timeliness (%) and likelihood of presentation for appointment (aOR). **Both coverage and timeliness.** | Pentavalent dose coverage.<br>**Group A** (Phone call reminders) = 100% coverage for all 3x Pentavalent doses.<br>**Group B** ('SMS-only' reminders) = 100% coverage for all 3x Pentavalent doses.<br>**Group C** ('SMS-Plus' health education SMS) = 100% for Penta1 and Penta2. 134 (99.2%) for Penta3.<br>**Group D** (Control), Penta1 = 131 (96.3%), Penta2 = 129 (94.9%), Penta3 = 127 (93.4%). | Timeliness of receipt of pentavalent doses (appropriate timing defined as 3days +/- scheduled appointment).<br>**Penta1: Group A** = 132 (99.2%), **Group B** = 132 (97.1%). **Group C** = 133 (98.5%). **Group D** = 71 (54.1%), (Chi-square 175.643, $p < 0.001^*$).<br>**Penta2: Group A** = 124 (93.2%). **Group B** = 109 (80.1%). **Group C** = 117 (86.7%). **Group D** = 93 (72.1%), (Chi 22.935 $p < 0.0001^*$). Only **Group A** had significant OR for Penta2 5.33 (95% CI 2.45-11.62, $p < 0.001^*$).<br>**Penta3: Group A** = 116 (87.2%). **Group B** = 95 (69.9%). **Group C** = 87 (64.9%). **Group D** = 85 (66.9%), (Chi 20.637 $p < 0.001^*$). Only **Group A** had significant OR for Penta3 3.37 (95% CI 1.8-6.33, $p < 0.001^*$). | Mother-infant pairs in all 3x intervention arms had significantly higher likelihood of presenting for subsequent immunisation visits than those in control group.<br>**Group A:** aOR 35.4 (95% CI 4.83-259.56, $p < 0.001^*$).<br>**Group B:** aOR 36.2 (4.94-265.42, $p < 0.001^*$).<br>**Group C:** aOR 7.133 (2.77-18.38, $p < 0.001^*$). | **Coverage:**<br>**(A)** LogOR 2.99 (95% CI 0.14-5.84)<br>**(B)** LogOR 3.01 (95% CI 0.16-5.87)<br>**(C)** LogOR 2.25 (95% CI 0.17-4.33)<br>**Timeliness:**<br>**(A)** LogOR 1.41 (95% CI 0.79-2.03)<br>**(B)** LogOR 0.33 (95% CI -0.18-0.83)<br>**(C)** LogOR 0.08 (95% CI -0.41-0.58) | The results indicate that phone call reminders are associated with the highest appropriateness of presentation timing for vaccinations. All 3x mHealth intervention groups were more likely to present for subsequent appointments, therefore these interventions might be more useful for immunisations requiring multiple doses. * = statistical significance ($p < 0.05$). |
| Kawakatsu *et al*, 2020 [62]. | Return rate for subsequent immunisation appointment (% and aOR). **Categorised as timeliness.** | Unadjusted return rate for subsequent immunisation appointment.<br>**Intervention:** 67% (95% CI 66%-68%).<br>**Control:** 62% (95% CI 61% - 64%) ($p < 0.001^*$).<br>aOR 1.17 (95% CI 1.05-1.31, $p < 0.05^*$) for return visits. | Number of return visits (subsequent appointments) on original appointment date.<br>**Intervention:** 2038 (41.7%).<br>**Control:** 1228 (35.7%) ($p < 0.001^*$). | N/A | **Coverage:** N/A<br>**Timeliness:** LogOR 0.25 (95% CI 0.16-0.34) | The results show that sending SMS text reminder is more likely to increase return visit rate. Mothers/caregivers are also more likely to show up for return visits and stick to original scheduled appointment dates, indicating the benefits for schedule adherence. * = statistical significance ($p < 0.05$). |
| Oladepo *et al*, 2020 [63]. | Completion of all immunisations (%) and coverage of Penta1, Penta2 and Penta3 (%). **Categorised as coverage.** | All immunisation completion rate.<br>**Intervention:** 76%<br>**Control:** 73.3%, ($p = 0.00^*$).<br>Pentavalent Dose Coverage:<br>**Penta1:** Intervention = 1082 (86.9%), Control = 938 (80.7%), ($p = 0.00^*$).<br>**Penta2:** Intervention = 1076 (87.5%), Control = 856 (74.8%), ($p = 0.00^*$).<br>**Penta3:** Intervention = 1035 (85%), Control = 799 (70.6%), ($p = 0.00^*$). | N/A | N/A | **Coverage:** LogOR 0.86 (95% CI 0.65-1.06)<br>**Timeliness:** N/A | The results show that there is a significant association between the intervention and completion of the 3x pentavalent vaccine doses. This can be attributable to the reminder messages intervention. * = statistically significant association (Chi-square). |
| Sampson *et al*, 2023 [64]. | Timeliness of immunisation rate (%) and reduction in dropout rate (%). **Categorised as timeliness.** | N/A | Immunisation timeliness rate.<br>**Intervention:** Baseline timeliness = 35% (289/827), Endline timeliness = 69% (522/757). Intervention effective at improving immunisation timeliness.<br>**Control:** Baseline timeliness = 64% (104/162), Endline timeliness = 67% (127/190). No statistical tests were conducted. | 60% reduction in dropout rate was recorded in intervention group between baseline and endline, and a 21% reduction in dropout was registered in the control group. | **Coverage:** N/A<br>**Timeliness:** LogOR 0.10 (95% CI 0.24-0.44) | Immunisation alert wristband intervention was shown to be effective at improving immunisation timeliness and reducing dropouts during immunisation schedule. It is certainly an interesting intervention that shows promise for mothers/caregivers with lower literacy and digital literacy. |
| Schlumberger *et al*, 2015 [65]. | Penta1, Penta2 and Penta3 coverage rate (%) and immunisation timeliness rate (%). **Both coverage and timeliness.** | Pentavalent dose coverage.<br>**Penta1:** Intervention = 187 (73.3%), Control = 150 (55.7%), (p<0.001*).<br>**Penta2:** Intervention = 182 (71.3%), Control = 144 (53.6%), (p<0.001*).<br>**Penta3:** Intervention = 165 (60.3%), Control = 126 (42.3%), (p<0.001*). | Pentavalent dose timeliness.<br>**Penta1:** Intervention = 113 (60.4%), Control = 73 (48.7%), (p=0.03*).<br>**Penta2:** Intervention = 81 (60.4%), Control = 49 (48.7%), (p=0.02*).<br>**Penta3:** Intervention = 45 (29.2%), Control = 29 (25.4%), (p=0.49). | N/A | **Coverage:**<br>LogOR 0.75 (95% CI 0.4-1.10)<br>**Timeliness:**<br>LogOR 0.58 (95% CI 0.08-1.08) | The results show that the SMS intervention was effective at increasing coverage of all 3x Penta doses in Burkina Faso context in comparison to control. The intervention led to significant improvement in increasing Penta1 and Penta2 timeliness, however, no significant difference was associated with Penta3 timeliness. * = statistical significance (p<0.05). |
| Yunusa *et al*, 2022 [66]. | Penta1, Penta2 and Penta3 coverage rate (%) and OR). **Categorised as coverage.** | Pentavalent dose coverage.<br>**Penta1:** Intervention = 210 (77.4%), Control = 194 (71.9%), OR 1.349 (95% CI 0.914-1.991, p=0.132).<br>**Penta2:** Intervention = 185 (68.3%), Control = 132 (48.9%), OR 2.249 (85% CI 1.585-3.191, p<0.0001*).<br>**Penta3:** Intervention = 163 (60.1%), Control = 117 (43.3%), OR 1.974 (95% CI 1.402-2.779, p<0.0001*).<br>Completion of all 3x Penta doses:<br>Intervention = 161 (59.4%), Control 92 (34.1%), OR 2.832 (95% CI 1.997-4.016, p<0.0001*). | N/A | Recommended that researchers use the local languages in sending the reminders to parents to enhance understanding. | **Coverage:**<br>LogOR 0.68 (95% CI 0.34-1.02)<br>**Timeliness:**<br>N/A | The results show that SMS combine with phone call immunisation reminders are effective in improving the completeness of all 3x pentavalent vaccine immunisation in the studied population, especially over time with the 2nd and 3rd doses. * = statistical significance (p<0.05). |
| Yunusa *et al*, 2024 [67]. | Penta1, Penta 2 and Penta3 coverage (%) and Timeliness of immunisation (%). **Both coverage and timeliness.** | Pentavalent dose coverage.<br>**Penta1:** Intervention = 196 (71.3%), Control = 133 (50.9%), (p<0.001*).<br>**Penta2:** Intervention = 175 (63.6%), Control = 75 (28.7%), (p<0.001*).<br>**Penta3:** Intervention = 169 (61.5%), Control = 44 (16.9%), (p<0.001*). | Pentavalent dose timeliness (receipt of immunisation within a month from the due date of recommended schedule).<br>**Penta1:** Intervention = 170 (61.8%), Control = 70 (26.8%), (p<0.001*).<br>**Penta2:** Intervention = 149 (54.2%), Control = 37 (14.2%), (p<0.001*).<br>**Penta3:** Intervention = 145 (52.7%), Control = 20 (7.7%), (p<0.001*). | N/A | **Coverage:**<br>LogOR 2.06 (95% CI 1.66-2.47)<br>**Timeliness:**<br>LogOR 2.60 (95% CI 2.08-3.11) | The results show that combined SMS and phone call reminders are effective in improving the completeness and timeliness of all 3x pentavalent doses for childhood immunisation in Nigeria. Article is in press. * = statistical significance (p<0.05). |

Note Penta1, Penta2 and Penta3 refers to 1st, 2nd and 3rd Pentavalent dose respectively, and DTP3 refers to third DTP dose. 95% CI = 95% Confidence Intervals, RR = Risk Ratio, OR = Odds Ratio, aOR = adjusted Odds Ratio, KES = Kenyan Shillings, BCG = Bacillus Calmette-Guérin vaccine, DTP = Diphtheria, Tetanus, and Pertussis vaccine, MCV = Measles-containing vaccine, OPV = Oral polio vaccine.

### Risk of Bias Assessments

Figure 3. shows the results of the RoB2 and ROBINS-I risk of bias assessments. Three of the included RCTs were considered as ‘Low risk’ [56, 57, 60]. All three of these studies were categorised as reporting outcomes relating to vaccination coverage, suggesting the findings relating to this outcome may have greater internal validity. Four were scored as having ‘Some concerns’, reporting on a mix of coverage, timeliness or both outcomes [55, 58, 59, 61]. Three were considered at ‘High risk’ [54, 65, 67]. Amongst the included non-RCTs, none were considered ‘Low risk’, two were scored as having ‘Moderate risk’ [61, 66], and two were considered as having ‘Critical risk’ of bias [63, 64]. This spread of risk of bias ratings highlights substantial variability in study quality, particularly among non-RCTs. The concentration of lower risk of bias studies reporting on vaccination coverage suggests more reliable conclusions can be made for this outcome. In contrast, caution is warranted when interpreting findings from studies assessed as having high or critical risk of bias (reported on both outcomes of interest). The presence of multiple studies with ‘some concerns’ or higher further emphasises the need for more rigorous, high-quality research to strengthen the evidence base.

**Figure 3.**
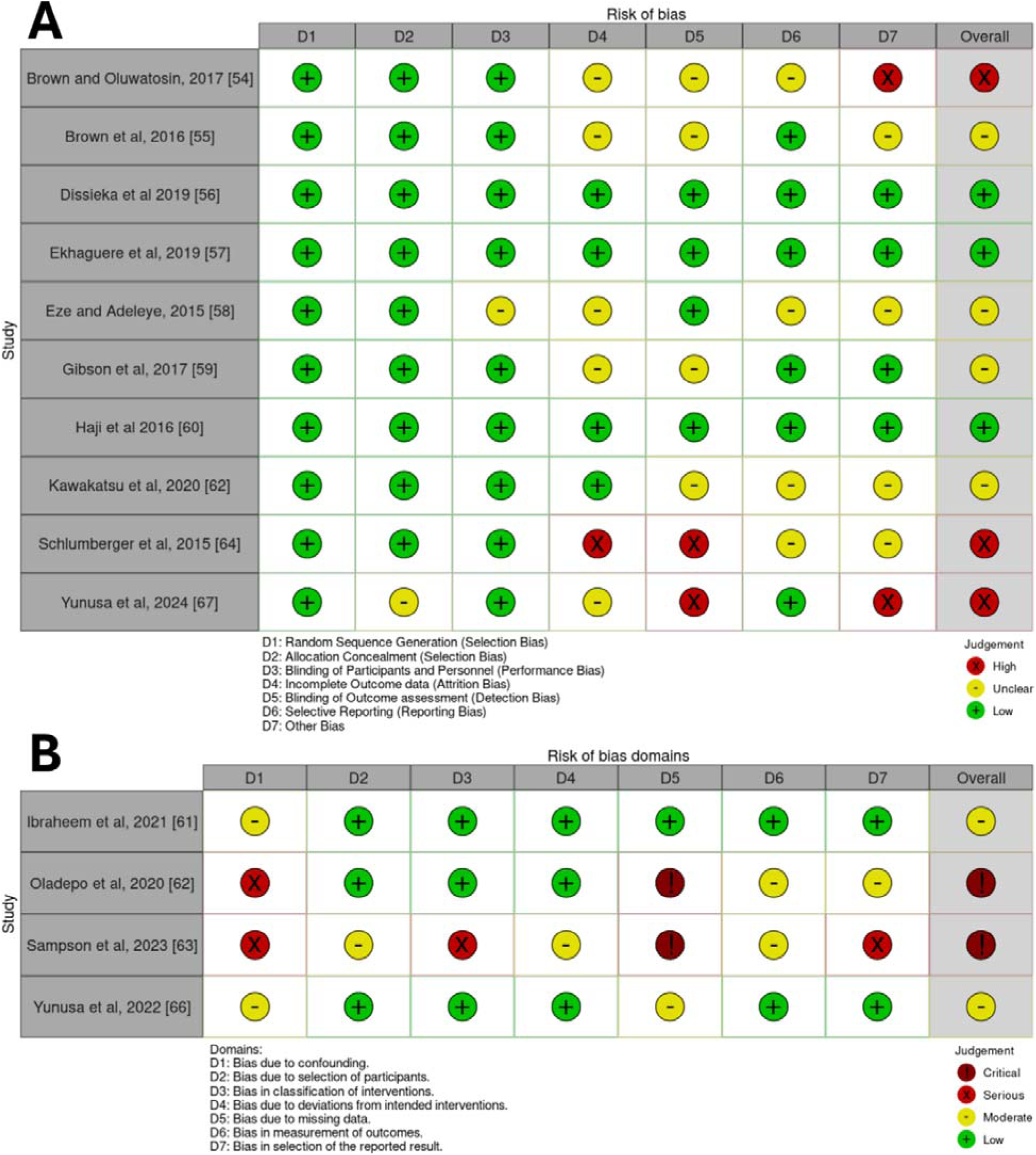
The risk of bias assessments of the included studies. (A) Risk of bias assessment of the RCTs using RoB2 [47], note that D7 ‘Other biases’ column refers to assessments of other important biases not assessed by RoB2, including assessing study population baseline imbalances, study design appropriateness, potential early stoppage of trials and potential conflicts of interest or funding biases (B) Risk of bias assessments of the non-RCTs using ROBINS-I [48].

### Study Objective 1 Findings – mHealth/DH intervention effect on vaccination coverage

Figure 4. provides a visual representation of the mHealth/DH interventions effectiveness in improving vaccination coverage of DTP/Pentavalent3 by study design.

**Figure 4.**
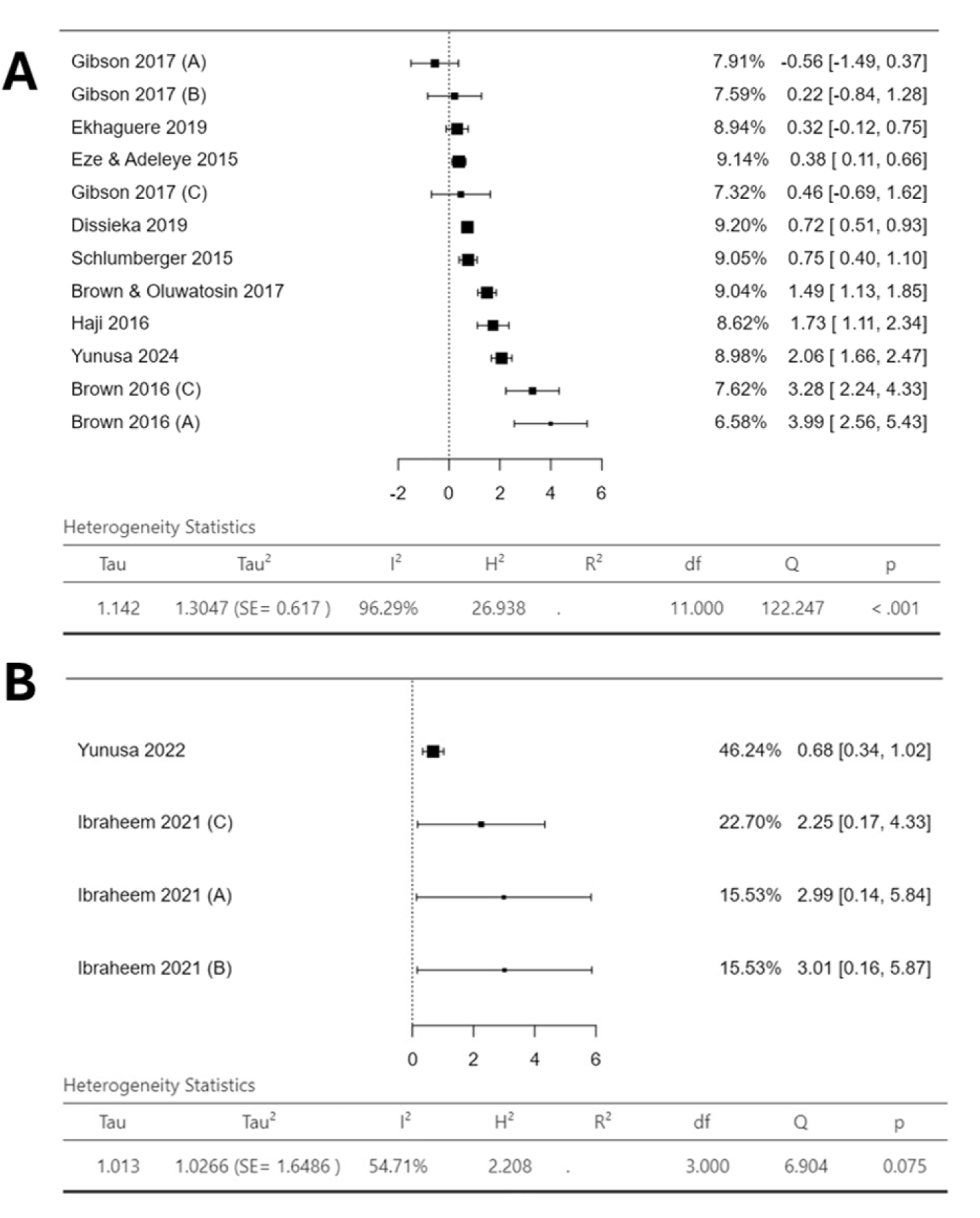
Forest plots showing logOR for mHealth/DH intervention effectiveness for increasing vaccination coverage of DTP/Pentavalent3. **(A)** logOR point estimates for the included RCTs **(B)** logOR point estimates for included non-RCTs*. % weighting of each study is shown which correlates to heterogeneity statistics along with 95% confidence intervals of the point estimates. As each intervention was analysed individually ‘Gibson 2017 (A)’ relates to Group A’s intervention from Gibson *et al* (2017) [59]. *Note Oladepo *et al* (2020) [61] has been excluded as an outlier as it had a weighting of 72.98% which resulted in masked heterogeneity (I^2^ = 0%). This large influence in the plot diminishes variability between studies.

Figure 4A shows the outcomes from the included RCTs. Extremely high heterogeneity was reported (I^2^=96.29%). Eleven interventions from the RCTs demonstrated a positive association between intervention and vaccination coverage. Only ‘Gibson 2017 (A)’, reported a negative outcome (-0.56 logOR, 95% CI -1.49 to 0.37) [59]. However, the confidence interval crosses the null (logOR = 0), indicating potential for a positive association. There are two outliers (which likely contribute to the elevated I^2^ statistic) displaying notably high effect estimates: ‘Brown 2016 (A)’ (3.99 logOR, 95% CI 2.56 to 5.43) and ‘Brown 2016 (C)’ (3.28 logOR 95% CI 2.24 to 4.33), both are phone call reminder interventions [55]. The remaining point estimates show a more homogeneous range, from 0.22 logOR (95% CI -0.84 to 1.28) to 2.06 logOR (95% CI 1.66 to 2.47). Overall, these findings indicate a likely positive association between mHealth/DH interventions and vaccination coverage.

Figure 4B shows associations between mHealth/DH interventions and vaccination coverage amongst the included non-RCTs. All four interventions presented here show positive logOR point estimates, suggesting these interventions were likely associated with increased vaccination coverage. Oladepo *et al* (2020) reported 0.86 logOR (95% CI 0.65-1.06) but was not included in the forest plot as its large weighting (due to sample size) masks heterogeneity (I^2^ = 0%), its overwhelming influence diminishes variability between studies [63]. With this excluded Figure 4B still displays moderate-high heterogeneity (I^2^ = 54.71%), thus grounds for narrative synthesis still stands.

### Study Objective 2 Findings – mHealth/DH intervention effect on vaccination timeliness

Figure 5 provides visual representation of the mHealth/DH intervention effect on timeliness of administration of DTP/Pentavalent3 by study design.

**Figure 5.**
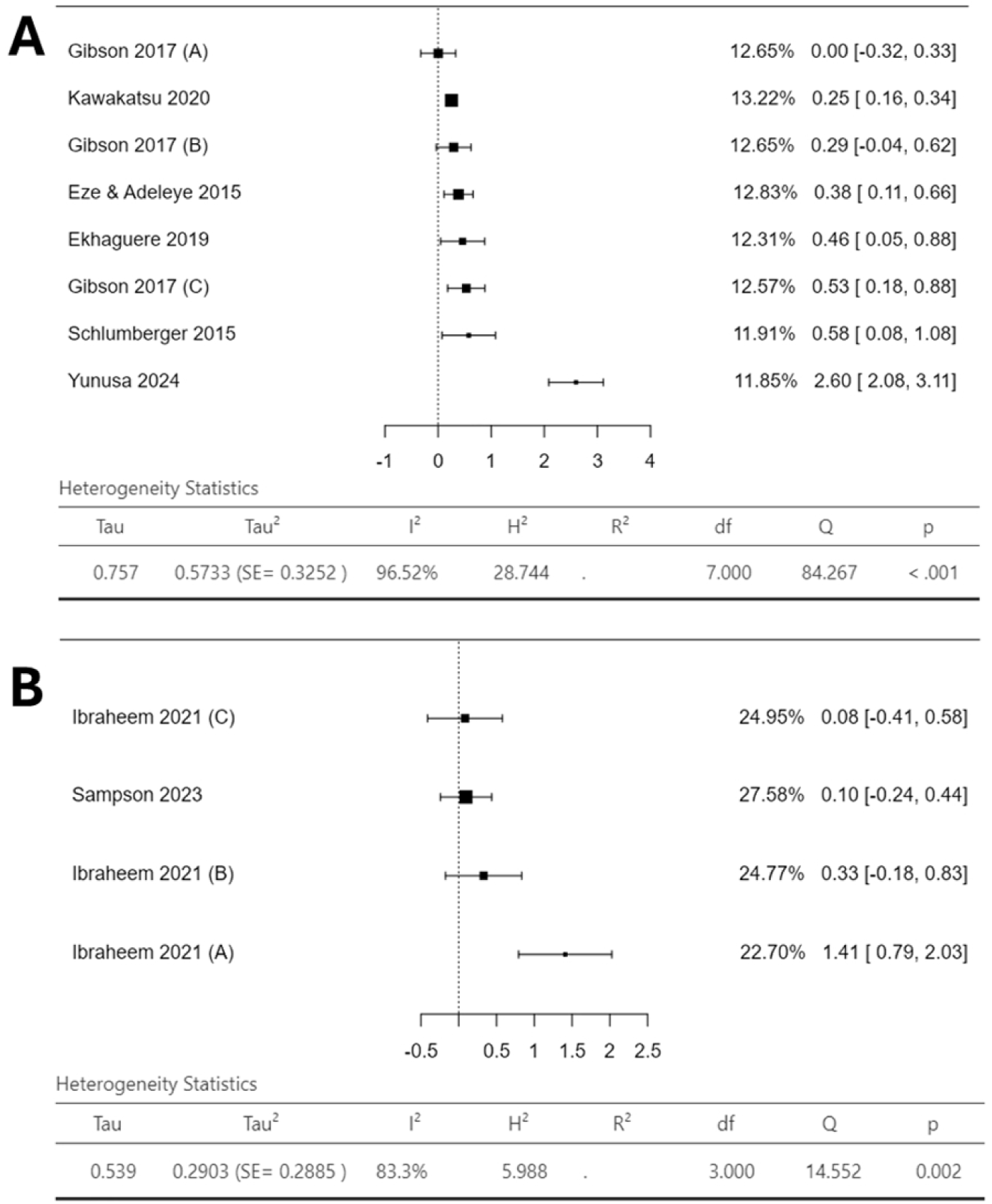
Forest plots showing logOR for mHealth/DH intervention effectiveness for improving timeliness of administration of DTP/Pentavalent3. (A) logOR point estimates for the included RCTs (B) logOR point estimates for included non-RCTs. % weighting of each study is shown which correlates to heterogeneity statistics along with 95% confidence intervals of the point estimates.

Figure 5A illustrates the interventions effect on timeliness from the included RCTs. Again, an extremely high I^2^ is reported (96.52%). These interventions showed fairly homogenous results, ranging from 0.00 logOR (95% CI -0.32 to 0.33) to 0.58 logOR (95% CI 0.08 to 1.08), indicating a positive association between mHealth/DH intervention and DTP/Pentavalent3 timeliness. Yunusa *et al* (2024) was a sole outlier (SMS and Phone call reminder intervention), reporting enhanced point estimate of 2.60 logOR (95% CI 2.08 to 3.11) [67].

Figure 5B shows the interventions investigated amongst the non-RCTs effect on timeliness. A high I^2^ is also reported (83.3%). Group B and C of Ibraheem *et al* (2021) and Sampson et al (2023), show fairly homogenous point estimates ranging from 0.08 logOR (95% CI -0.41 to 0.58) to 0.33 logOR (95% CI -0.18 to 0.83) [61, 64]. The only outlier was Group A of Ibraheem *et al* (2021), showing 1.41 logOR (95% CI 0.79 to 2.03) [61].

### Study Objective 3 Findings - Additional results relevant to mHealth/DH intervention implementation

Two studies provided useful additional information [56, 66]. Dissieka *et al* (2019) reported that when given a choice between receiving SMS or voice message appointment reminders, 675 (84.6%) of mothers/caregivers chose to receive voice message reminders, whereas only 123 (15.4%) chose to receive SMS messages, and voice message reminders were preferred by 97.7% in rural areas [56]. Additionally, Yunusa *et al* (2022) recommended sending SMS messages in local languages, and to not assume English is understood, an important consideration for future intervention development [66].

### GRADE Certainty of Evidence Assessment

The GRADE assessment was conducted separately around each of the intervention-type subgroups and the two primary outcome groups (vaccination coverage and vaccination timeliness). **Table 3** shows the GRADE summary of findings table. The full explanations for GRADE scoring are shown in **Supplementary Information** (**S5**). In the vaccination coverage outcome group, the findings from ‘SMS-only’ and ‘SMS reminders and/or Voice messages or Phone calls’ subgroups were scored as having ‘Moderate’ certainty. The ‘Phone call-only’ subgroup scored ‘Low’ certainty, whilst the ‘SMS-Plus’ subgroup’s findings were considered ‘Very low’ certainty. For the vaccination timeliness findings, the ‘SMS-only’ and ‘SMS-Plus’ intervention subgroups’ findings were considered as having ‘Moderate’ certainty, whereas the ‘SMS reminders and/or Voice messages or Phone calls’, ‘Phone call-only’ and ‘Electronic Immunisation Alert Wristband’ all scored ‘Very low’ certainty.

**Table 3.** GRADE Summary of findings table showing certainty of evidence for each outcome and intervention-type pairings.

| Outcome | Intervention Type Subgroup | Study Design (RCT or Non-RCT) | Risk of Bias | Inconsistency | Indirectness | Imprecision | Publication Bias | Effect Estimate Range (LogOR (95% CI)) | Certainty of Evidence (GRADE) |
| --- | --- | --- | --- | --- | --- | --- | --- | --- | --- |
| Vaccination Coverage | SMS-only | Both, RCT (n=4) & Non-RCT (n=2) | Serious Concerns | No Serious Concerns | No Serious Concerns | No Serious Concerns | No Serious Concerns | -0.56 LogOR (95% CI -1.49-0.37) to 3.01 LogOR (95% CI 0.16-5.87) | Moderate ⊗⊗⊗ |
|  | SMS-Plus | Both, RCT (n=2) & Non-RCT (n=2) | Very Serious Concerns | Serious Concerns | Serious Concerns | Very Serious Concerns | No Serious Concerns | 0.22 LogOR (95% CI -0.84-1.28) to 2.25 LogOR (95% CI 0.17-4.33) | Very Low ⊗ |
|  | SMS reminders and/or Voice messages or Phone calls | RCT (n=3) | Serious Concerns | No Serious Concerns | No Serious Concerns | No Serious Concerns | No Serious Concerns | 0.32 LogOR (95% CI -0.12-0.75) to 2.06 LogOR (95% CI 1.66-2.47) | Moderate ⊗⊗⊗ |
|  | Phone call-only | Both, RCT (n=3) & Non-RCT (n=1) | Serious Concerns | No Serious Concerns | No Serious Concerns | Some Concerns | Serious Concerns | 1.49 LogOR (95% CI 1.13-1.85) to 3.99 LogOR (95% CI 2.56-5.43) | Low ⊗⊗ |
| Vaccination Timeliness | SMS-only | Both, RCT (n=3) & Non-RCT (n=1) | No Serious Concerns | No Serious Concerns | No Serious Concerns | Serious Concerns | No Serious Concerns | 0.00 LogOR (95% CI -0.32-0.33) to 0.38 LogOR (95% CI 0.11-0.66) | Moderate ⊗⊗⊗ |
|  | SMS-Plus | Both, RCT (n=2) & Non-RCT (n=1) | No Serious Concerns | No Serious Concerns | Serious Concerns | No Serious Concerns | No Serious Concerns | 0.08 LogOR (95% CI -0.41-0.58) to 0.53 LogOR (95% CI 0.18-0.88) | Moderate ⊗⊗⊗ |
|  | SMS reminders and/or Voice messages or Phone calls | RCT (n=2) | Serious Concerns | Serious Concerns | No Serious Concerns | Some Concerns | Serious Concerns | 0.46 LogOR (95% CI 0.05-0.88) to 2.60 LogOR (95% CI 2.08-3.11) | Very Low ⊗ |
|  | Phone call-only | Non-RCT (n=1) | No Serious Concerns | No Serious Concerns | No Serious Concerns | Serious Concerns | Serious Concerns | 1.41 LogOR (95% CI 0.79-2.03) | Very Low ⊗ |
|  | Electronic Wristband | RCT (n=1) | Very Serious Concerns | No Serious Concerns | No Serious Concerns | Serious Concerns | Serious Concerns | 0.10 LogOR (95% CI -0.24-0.44) | Very Low ⊗ |

\*See **Supplementary Information (S5)** for full explanations on scoring domains.

### Narrative synthesis

For the narrative synthesis the studies have been grouped by mHealth/DH intervention-type. The description and which studies included in each subgroup is shown in **Supplementary Information** (**S4**).

### Study Objective 1 Narrative Synthesis – mHealth/DH intervention effect on vaccination coverage

Four mHealth/DH intervention-type subgroups reported outcomes categorised as vaccination coverage (‘SMS-only’ appointment reminders, ‘SMS-Plus’, ‘SMS reminders and/or Voice messages or Phone calls’, and ‘Phone call-only’ reminders).

#### (i) ‘SMS-only’ appointment reminders (including automated and non-automated)

Five ‘SMS-only’ interventions showed a positive association with vaccination coverage [58, 60, 61, 65, 66]. Group A in Gibson *et al* (2017) reported a negative logOR (-0.56 (95% CI - 1.49 to 0.37)) [59], however, as the confidence interval crosses the null, a positive association might be possible. Contrastingly, Group B of Ibraheem *et al* (2021) demonstrated a much greater point estimate of 3.01 logOR (95% CI 0.16 to 5.87) [61]. However, the broad confidence interval range suggests imprecision in the point estimate, which may limit the reliability of this finding. The remaining four studies all showed a positive association between ‘SMS-only’ reminders and vaccination coverage, ranging from 0.38 logOR (95% CI 0.11 to 0.66) to 1.71 logOR (95% CI 1.11 to 2.34) [58, 60, 65, 66]. The intervention-outcome group pairing scored moderate certainty of evidence; therefore, we have moderate confidence in these findings and recognise that further research may lead to alternative conclusions.

Overall, the results suggest that implementing ‘SMS-only’ appointment reminders during the malaria vaccine rollout is preferable to no intervention but may lead to only modest positive effects on increasing vaccination coverage. It is likely that other types of mHealth/DH interventions will be more effective for improving vaccination coverage. For ‘SMS-only’ interventions to be effective, mothers/caregivers must receive the SMS reminder [68]. If a spouse or relative’s phone is used, then this opens the possibility of reminders being missed, however, it was unclear whether SMS reminders were received. As the SMS messages must be read and understood by recipients to be effective, the modest results exhibited could be explained because of lower literacy rates amongst recipients and may be more prominent in rural communities where literacy rates might be lower [69, 70].

#### ((ii) ‘SMS-Plus’ (SMS appointment reminders plus cash incentives or health education SMS messages)

The ‘SMS-Plus’ interventions show great variation in logOR, likely due to the diversity of intervention enhancements. Oladepo *et al* (2020) reported the most precise point estimate, 0.86 logOR (95% CI 0.65 to 1.06), however, due to potential limitations identified in risk of bias assessment, we acknowledge this finding’s reliability may be reduced. [63]. Group B and C of Gibson *et al* (2017), which combined SMS reminders with cash incentives, reported 0.22 logOR (95% CI -0.84 to 1.28) and 0.46 logOR (95% CI -0.69 to 1.62), respectively [59]. Confidence intervals in both crossed the null, reflecting great uncertainty in point estimate precision. Group C of Ibraheem *et al* (2021), showed 2.25 logOR (95% CI 0.17 to 4.33), with a wide confidence interval range, again indicating extreme uncertainty in point estimate precision [61]. Overall, the certainty of evidence for this intervention-outcome pairing was very low, which limits the strength of conclusions regarding the effectiveness of ‘SMS-Plus’ interventions for increasing vaccination coverage

The findings suggest that implementing a ‘SMS-Plus’ intervention may modestly improve vaccination coverage, however the imprecision of the estimates means there is possibility the interventions will have little or no effect. An interesting observation was that health education intervention reported by Oladepo *et al* (2020), showed greater effect in comparison to cash incentivisation interventions for vaccination coverage [61, 63]. As ‘SMS-Plus’ interventions are also SMS-based, they also require SMS receipt and recipient understanding of SMS contents to be effective which could explain the similar outcomes as ‘SMS-only’ interventions. As interventions were ‘enhanced’ through cash incentivisation or health educational messaging, stronger associations between interventions and outcomes were expected, however, were not the found. Further context-specific study on whether these intervention-types will offer value despite utilising extra resources is required.

#### (iii) ‘SMS reminders and/or Voice messages or Phone calls’

All three studies in this subgroup showed positive associations between intervention and vaccination coverage with varying degrees of precision [56, 57, 67]. Yunusa *et al* (2024) reported the strongest association, logOR of 2.06 (95% CI 0.17 to 4.33), however, this finding’s validity may be influenced by the risk of bias assessment and should be interpreted accordingly [67]. Ekhaguere *et al*’s (2019) intervention (combining SMS and voice messages), reported a positive 0.32 logOR (95% CI -0.12 to 0.75), however, as the confidence interval range crosses the null there is uncertainty around the estimate’s precision [57]. Dissieka *et al* (2019) carried the most weight when considering risk of bias assessment, reporting 0.72 logOR (95% CI 0.51 to 0.93) [56]. The point estimate suggests that providing mothers/caregivers with a choice of SMS or voice messages is positively associated with increased vaccination coverage. For this intervention-outcome group, the certainty of evidence was again moderate, indicating sufficient confidence that these interventions will positively affect vaccination coverage.

Overall, the interventions in this subgroup showed robust positive associations, suggesting that combining SMS reminders with voice messages or phone calls, or giving mothers/caregivers the choice between SMS reminders or voice-based reminders would be effective and benefit vaccination coverage. This could be due to voice-based interventions negating the limitation of literacy. Furthermore, offering mothers a choice of intervention, may make them feel more empowered in relation to their child’s health [71]. The moderate certainty of evidence indicates reasonable confidence in this finding. Based on the current evidence, these findings are sufficient to inform recommendations for the malaria vaccine rollout. Given that these interventions are potentially more resource-intensive, further research evaluating their cost-effectiveness is imperative.

#### (iv) ‘Phone call-only’ reminders

All three interventions involving phone call reminders show great positive associations for vaccination coverage [54, 55, 61]. All point estimates were >1. Brown & Oluwatosin (2017) provided the most precise estimate in the subgroup, with logOR of 1.49 (95% CI, 1.13 to 1.85) [54]. Group A in Ibraheem *et al* (2021), reported 2.99 logOR (95% CI, 0.14 to 5.84), however the wide confidence interval range indicates substantial uncertainty in this point estimate’s precision, raising concerns around its reliability [61]. Group A of Brown *et al* (2016), reported highest logOR of 3.99 (95% CI, 2.56 to 5.43) [55]. Although the GRADE assessment rated this intervention-outcome group as having low certainty of evidence, the point estimates were consistently greater than those of other intervention-types, suggesting that phone call reminder interventions may offer the greatest potential for increasing vaccination coverage. While these findings support the hypothesis that interventions incorporating voice-based components may be more effective and certainly show promise, the low certainty of evidence limits their immediate applicability for informing public health policy and practice. Nonetheless, these findings highlight the potential of phone-call reminders for improving vaccination coverage and point to a clear need for further research evaluating these potentially effective interventions.

### Study Objective 2 Narrative Synthesis – mHealth/DH intervention effect on vaccination timeliness

Five mHealth/DH intervention-type subgroups reported outcomes categorised as vaccination timeliness (‘SMS-only’ appointment reminders, ‘SMS-Plus’, ‘SMS reminders and/or Voice messages or Phone calls’, ‘Phone call-only’ reminders, and ‘Electronic immunisation alert wristband’).

#### (i) ‘SMS-only’ appointment reminders (including automated and non-automated)

Three ‘SMS-only’ interventions showed positive associations between intervention and vaccination timeliness [58, 61, 62]. Group A of Gibson *et al* (2017), showed no effect, 0.00 logOR (95% CI -0.32 to 0.33), however, the wide confidence interval range, indicates uncertainty around true point estimate [59]. The remaining point estimates were fairly homogeneous showing slight positive associations, with Kawakatsu *et al* (2020) reporting 0.25 logOR (95% CI 0.16 to 0.34) [62], and Group B of Ibraheem *et al* (2021) reporting 0.33 logOR (95% CI -0.18 to 0.83). Although they reported a positive association, the wide confidence interval range crosses the null, indicates potential for no effect [61]. Eze and Adeleye (2015) also reported a slight positive association of 0.38 (95% CI 0.11 to 0.66) [58]. A moderate certainty of evidence was found for this intervention-outcome pairing. Like the vaccination coverage results, these findings suggest that ‘SMS-only’ interventions are likely to be more effective than no intervention but may only lead to modest improvements in vaccination timeliness. The potential reasons underlying the coverage findings are also applicable here.

#### (ii) ‘SMS-Plus’ (SMS appointment plus cash incentives or health education SMS)

All three interventions showed positive associations between intervention and timeliness [59, 61]. Group C of Ibraheem *et al* (2021), received health education SMS’, reporting only a slight positive association of 0.08 logOR (95% CI -0.41 to 0.58) [61], when considering the wide confidence interval range there is possibility of no effect. Groups B and C of Gibson *et al* (2017) combined SMS appointment reminders with cash incentives of 75KES and 200KES respectively [59]: Group B reported 0.29 logOR (95% CI -0.04 to 0.62), however, again the confidence interval crossed the null, whilst Group C demonstrated a stronger positive association of 0.53 logOR (95% CI 0.18 to 0.88). Again, these findings were assessed as having moderate certainty, indicating satisfactory confidence. The findings suggest that combining SMS appointment reminders with larger cash incentives may have greater effect for improving vaccination timeliness. However, further research, particularly cost-effectiveness analyses, is warranted to support decisions on enhancing interventions for increasing vaccination timeliness.

#### (iii) ‘SMS reminders and/or Voice messages or Phone calls’

Only two interventions in this subgroup reported timeliness outcome data [57, 67]. Yunusa *et al* (2024) reported a high logOR of 2.60 (95% CI 2.08 to 3.11) [67]. Whereas Ekhaguere *et al*’s (2019) intervention combining SMS and voice messages showed a logOR of 0.46 (95% CI, 0.05 to 0.88) [57]. This intervention-outcome group was assessed as having very low certainty of evidence, indicating uncertainty in the findings.

Overall, the findings suggest that combining SMS reminders with a voice-based component may have a positive effect on vaccination timeliness, supporting the hypothesis that voice-based interventions could be more effective by mitigating some of the limitations associated with ‘read-only’ SMS interventions. However, these findings should be interpreted with caution given the very low certainty of evidence.

#### (iv) ‘Phone call-only’ reminders

Group A of Ibraheem *et al* (2021) was the sole ‘phone call-only’ reminder intervention showing effect on timeliness, reporting logOR of 1.41 (95% CI, 0.79 to 2.03) [61]. It’s relatively narrow confidence intervals and satisfactory risk of bias, suggests that implementing phone call reminders would likely improve immunisation timeliness. Although, this is in line with the study findings, the very low certainty of evidence score means uncertainty around this finding must be acknowledged, and further research is required for more reliable conclusions.

#### (v) ‘Electronic immunisation alert wristband’

Only one study implemented the wearable electronic immunisation alert wristband reporting only a slight positive association, 0.10 logOR (95% CI, -0.24 to 0.44) [64]. Additionally, the confidence interval crossed the null, demonstrating potential for no effect. Again, a very low certainty of evidence score was registered.

## Discussion

### Overview

This systematic review and subsequent narrative synthesis investigated which mHealth/DH interventions are most effective at increasing vaccination uptake (of DTP/Pentavalent vaccine) in the 19 SSA countries that were due to roll out malaria vaccine in 2024. Most studies were conducted in West Africa (10 from Nigeria [54, 55, 57, 58, 61-64, 66, 67], 1 from Burkina Faso [65], and 1 from Cote D’Ivoire [56]), making the findings more generalisable to that setting. The remaining two studies were conducted in Kenya, East Africa [59, 60]. The included mHealth/DH interventions were all related to communication technology and immunisation appointment reminders. The intervention-types investigated included: ‘SMS-only’ appointment reminders, ‘SMS-Plus’ appointment reminders, ‘SMS messages and/or voice messages or phone calls’, ‘phone call-only’ appointment reminders, and a wearable electronic immunisation alert wristband. Extremely high heterogeneity across the studies was reported, which seems to be common when investigating mHealth/DH interventions and was reported in similar reviews [34, 37]. Thus, a narrative synthesis was conducted by grouping mHealth/DH intervention-type and assessed their effectiveness for improving vaccination coverage and timeliness.

A positive association between intervention and improvement in vaccination coverage was reported for 16 interventions (Figure 3), and 11 interventions showed a positive association between intervention and vaccination timeliness of DTP/Pentavalent3 administration (Figure 4). This consistency in findings across the included studies, enhances the evidence’s reliability. However, since almost all reported interventions showed a positive association, concerns around publication bias must be acknowledged. Conducting a more rigorous grey literature search and searching of trial registries may have reduced this potential reporting publication bias and increase this review’s external validity [72]; however, this was out with this review’s scope. Despite this limitation, the review does provide some robust evidence for informing policy and practice on mHealth/DH intervention implementation in the malaria vaccine immunisation programmes.

### SMS-based interventions (‘SMS-only’ and ‘SMS-Plus’)

When considering intervention-type subgroups, the findings suggests that voice-based interventions (phone call or voice message reminders), are likely more effective than SMS-based interventions. Only slight positive associations were observed for both vaccination coverage and timeliness in ‘SMS-only’ appointment reminder interventions, with moderate certainty of evidence. This suggests that sending out ‘SMS-only’ appointment reminders is more favourable than no intervention, however, other types of mHealth/DH intervention are likely to be more effective for improving vaccination coverage and timeliness. The stronger ‘SMS-only’ positive associations were reported in urban areas, indicating potentially greater generalisability to these settings.

‘SMS-Plus’ interventions also showed positive associations, however, stronger associations in comparison to ‘SMS-only’ interventions were not found despite ‘SMS-Plus’ interventions being enhanced. The certainty of evidence for the effect of ‘SMS-Plus’ interventions on vaccination coverage was very low, highlighting uncertainty in the findings. Whereas the ‘SMS-Plus’ and vaccination timeliness intervention-outcome group was assessed as having moderate certainty of evidence, indicating much greater reliability in findings for that outcome.

Interestingly, although some ‘SMS-Plus’ interventions offered mothers/caregivers cash incentives for attending immunisation appointments, it was the SMS health educational interventions that demonstrated stronger associations with improved vaccination coverage. When considering SMS reminders combined with cash incentives, offering a larger incentive (200KES = ∼1.2GBP), appeared to have greater effect on both vaccination coverage and timeliness compared to the lower amount (75KES = ∼0.45GBP) [59]. However, according to the GRADE assessment, the evidence supporting these interventions for improving vaccination timeliness was more reliable. In the wider literature, providing mothers with 2USD when their child received timely pentavalent vaccine doses, was found to increase the proportion of individuals receiving timely pentavalent doses [73]. However, this study utilised a small study population, lowering its validity. In contrast, another systematic review, specifically investigating financial incentives for increasing coverage of child health interventions, found limited effect on immunisation coverage when financial incentives were offered, indicating this may not be a useful intervention [74]. Interestingly, an RCT in Ghana found that offering small financial incentives to health care workers improved vaccination coverage and timeliness of BCG and OPV [75], suggesting that incentivising those administering vaccinations might be more effective.

A major limitation of both ‘SMS-only’ and ‘SMS-Plus’ interventions is the possibility that target recipients do not receive the SMS reminders. The ability to log SMS delivery was beyond the scope of the included studies, and assumptions were made on target population receiving SMS messages. This presents an opportunity for future research investigating mHealth SMS intervention delivery success rate in SSA context. Additionally, for SMS-based interventions to be successful, recipients must read and understand the respective messages, highlighting another potential limitation. Low literacy amongst mothers/caregivers could result in them not understanding the contents of SMS reminders, severely limiting their effectiveness. ‘SMS-Plus’ interventions involving sending educational messages around vaccination may be particularly susceptible to this limitation. This could be particularly prominent in rural communities where literacy rates might be lower. The intervention showing least effect in the ‘SMS-only’ subgroup was conducted in a rural area which supports this hypothesis, therefore is an important consideration [59]. The potential limitation around literacy was also reported in a SMS-based data collection study amongst midwives in Liberia, it concluded SMS interventions must be targeted towards those with higher literacy [76]. Although, a certain literacy level is required for these interventions to be effective, new mothers/caregivers should be encouraged to seek help if illiterate, and strategies to complement SMS-based interventions developed to address this barrier. SMS messages should be made simple and easy to understand, as highlighted in a study in rural Kenya [77]. Investigating the association between participant educational attainment and mother/caregiver literacy and SMS immunisation appointment reminder effectiveness, perhaps through an observational case-control study, would provide valuable insight into this potential limitation.

Despite the outlined limitations of SMS-based interventions, in contrast it could be argued that implementing basic SMS-based interventions increases accessibility and may have wider reach. Although SMS-based interventions have fewer capabilities (e.g. extremely limited multi-media content delivery and tracking capabilities), SMS-compatible devices are cheaper and are supported on basic ‘second generation’ 2G cellular connectivity networks which are much more widely available in rural areas [78]. Therefore, when considering connectivity and affordability barriers, implementing basic interventions may be more equitable.

The findings in this review and wider literature suggest that implementing ‘SMS-only’ interventions for the malaria vaccine rollout could be effective, therefore their development should be considered. Furthermore, SMS interventions have already been shown to be effective in the context of malaria control [79]. The findings show that enhancing SMS interventions (‘SMS-Plus’) with educational messages is likely to be more effective than cash incentives for improving vaccination coverage, whereas the addition of cash incentives appeared to have greater effect on improving vaccination timeliness, potentially by creating a sense of urgency. Larger cost-effectiveness analyses are required to determine whether the additional resources needed to implement enhanced interventions provide sufficient value.

### Voice-based interventions (‘SMS reminders and/or Voice messages or Phone calls’ and ‘Phone call-only’)

Again, all interventions in the ‘SMS reminders and/or Voice messages or Phone calls’ subgroup showed positive associations for both vaccination coverage and timeliness. The intervention showing the strongest association combined SMS and phone call reminders, suggesting that implementing combination would likely be effective for increasing vaccination coverage and timeliness [67]. The certainty of evidence for this intervention-type subgroup was assessed as moderate for vaccination coverage, and very low for vaccination timeliness, indicating that recommendations are more reliable for the vaccination coverage context. When considering risk of bias, Dissieka *et al*’s (2019) finding was considered the most reliable in the subgroup [56]. Their RCT was conducted in rural Cote D’Ivoire and offered participants the choice of receiving SMS or voice message reminders. Interestingly, most participants chose to receive voice messages, demonstrating that when given the choice, voice messages were the preferred intervention. This finding supports the hypothesis that effectiveness of SMS-based interventions is potentially limited in rural populations with lower literacy. Although Cote D’Ivoire does have relatively high literacy rates in comparison to other SSA nations, female literacy is lower, and as mothers/caregivers are the main target of these interventions [80]. This could explain the finding and further highlights the importance of considering target demographic literacy, particularly in rural settings when developing mHealth/DH interventions. It is acknowledged that only this one study offered participants a choice of intervention, thus limiting its generalisability. However, it does highlight another research gap and need for further investigation into providing choices of intervention in the immunisation programme context.

Using voice messages has been successful in other contexts in SSA, for example a Nigerian RCT demonstrated that women receiving voice messages were more likely to attend antenatal care appointments [81]. Furthermore, they were implemented in rural Senegal to improve infant and child feeding practices [82]. This certainly highlights the potential for voice-message interventions; however, we recognise that combining SMS interventions with voice-based components may be more resource-intensive. Again, further cost-effectiveness study would help clarify its viability. Overall, the findings demonstrate that these interventions are likely to be effective in the malaria vaccine context for increasing vaccination coverage, and offering participants the choice of intervention (SMS or voice message) should be considered during intervention development.

Generally, the positive associations reported in the ‘Phone call-only’ subgroup were stronger in comparison to the other subgroups. This further supports the hypothesis that interventions with voice-based components are more effective. This could be due to recipients responding to better to more engaging and personal interventions [83]. Implementing phone calls also open the opportunity for recipients to potentially seek clarification on queries relating to vaccination and may allow reinforcement messaging about importance of vaccination and adhering to outlined vaccination schedules by vaccinators. Furthermore, using phone calls would overcome the outlined limitations of SMS-based reminders, with verbal information more likely to be understood. A Cochrane review investigating various patient reminder interventions, mirrored this review’s findings reporting that phone call reminders were the most effective intervention [84]. However, they did raise the concern around phone call reminders being more costly than other methods. While interventions incorporating phone call reminders appear to be most effective in increasing vaccination coverage and timeliness and show promise, further study is required to increase the findings’ validity.

Despite the presented evidence showing this intervention is likely to be effective, cost-effectiveness comparison between SMS and phone calls within the malaria vaccine immunisation programme context must be conducted. Another systematic review reported that phone call reminders were effective, however, two of their included studies reported that costs per SMS reminder were considerably lower than phone calls, again highlighting this intervention’s potentially costly nature [85]. With expansion of AI and voice technology, the cost-barriers of these interventions may be reduced and should be strongly considered for future immunisation programmes following cost-effectiveness analysis [86].

### Other interventions

The final intervention subgroup was the ‘Electronic immunisation alert wristband’. Although an interesting intervention, only a very slight positive association was shown. Its imprecision and very low certainty of evidence mean recommendations cannot be made supporting this intervention. A similar intervention in Pakistan also showed limited effect [87].

### mHealth/DH intervention recommendation

Overall, our review’s findings show implementation of a mHealth/DH intervention in this specific context would likely be beneficial for the malaria vaccine rollout for increasing vaccination coverage and improving timeliness. A combination of SMS appointment reminders with either voice messages or phone calls would likely be the most effective intervention. Whilst this approach appears promising for improving vaccination coverage, questions remain regarding its effectiveness for enhancing vaccination timeliness.

### Findings in comparison to other systematic reviews

As previously mentioned, several similar systematic reviews were identified during scoping searches. Gilano *et al* (2024) investigated the effect of mHealth on all essential childhood immunisations. Despite reporting extremely high heterogeneity, they conducted a meta-analysis and reported an OR of 2.21 for Penta3 dose [37]. Our narrative findings mirror this and further support use of mHealth/DH interventions to increase vaccination coverage. Eze *et al* (2021) solely focused on SMS appointment reminders in LMICs in general and like our review focused on DTP/Pentavalent vaccine [34]. Again, despite reporting high heterogeneity, a meta-analysis found that implementing SMS reminders significantly improved childhood immunisation coverage (RR = 1.16, 95% CI 1.10 to 1.21) and timeliness (RR = 1.21, 95% CI 1.12 to 1.30) [34]. Our systematic review did find similar conclusions that mHealth/DH interventions of all types including SMS appointment reminders, were likely to improve vaccination coverage and timeliness. The authors recognised phone call reminders would be advantageous for reaching populations with limited or no education, and future studies should explore a combined intervention for optimising immunisation outcomes. This mirrors the ideas generated by in review, further increasing the evidence-base.

### Geographic generalisability of results in SSA

This review aimed to assimilate evidence of mHealth/DH interventions for increasing uptake of DTP/Pentavalent vaccine as predictors for malaria vaccines in the 19 SSA countries targeted to roll them out in 2024. However, studies from only 4 of these countries were included, meaning 15 were not represented, limiting the review’s generalisability. Most studies were from Nigeria, where the highest global malaria burden is reported [21, 88]. Therefore, implementing mHealth/DH interventions to support Nigeria’s malaria vaccine programmes could improve both vaccination coverage and timeliness, and potentially reduce Nigeria’s high malaria-attributed childhood mortality and morbidity. Studies have shown that implementing mHealth/DH interventions in Nigeria is likely to be acceptable [89-91].

However, low phone ownership and literacy rates have been shown in rural Nigerian communities [92], highlighting the need to consider these context-specific aspects.

Although 15 SSA countries were not represented in our review, there is evidence showing mHealth/DH interventions are being implemented in these countries. A comparative review on mHealth and eHealth use in SSA included studies from 17 SSA countries [93]. It reported Uganda, South Africa, Madagascar and Kenya as the SSA countries with the most mHealth/DH studies. Aboye *et al* (2023), also found Uganda to be most represented (along with Kenya and Nigeria) [6]. Whilst South Africa and Madagascar were out with this review’s scope, it is questionable why Uganda was not represented here. There is evidence of mHealth/DH use in Uganda in the context of vaccination, for example using DHIS2 [94]. Additionally, one of the excluded studies in this review was an RCT protocol on SMS to reduce vaccination dropouts in Uganda, showing research is happening there [95]. mHealth/DH interventions have also been used in more of the 15 SSA countries not represented here such as Ghana, Sierra Leone, and South Sudan [71, 96, 97]. This suggests that implementing a mHealth/DH intervention in these countries’ malaria vaccine immunisation programme is possible, however, it is acknowledged that country-specific messaging preferences should be explored.

### Impact and important considerations

It is well established that essential childhood vaccinations contribute hugely to public health preventing millions of deaths annually, and finding strategies to increase their uptake and coverage, whilst ensuring their timely administration is essential to maximise their protective benefits. Although many of the mHealth/DH interventions in this review showed only a small effect, when scaled to population level they could have great positive impact. As full rollout of the malaria vaccines began in 2024, there is huge potential to alleviate some of the malaria-attributed childhood mortality and morbidity, and the presented evidence in this review suggests that mHealth/DH interventions could support health systems and immunisation programmes in SSA to increase vaccine reach. However, country-specific studies on intervention acceptability and feasibility in both urban and rural settings, must be conducted to assess their effectiveness in specific contexts. Additionally, the findings highlight the need for considerations around target population literacy and technology accessibility, particularly in relation to implementing SMS-based interventions. Implementing voice-based interventions may overcome some of these barriers, however, questions around costs may limit feasibility in more resource limited settings. It must be acknowledged that accessibility and affordability of mobile devices remain significant barriers in LMICs, with many people lacking mobile phone access or sufficient digital literacy [98]. High mobile data connectivity costs in LMICs are also potentially prohibitive, which would limit these interventions [33, 99]. These challenges are arguably more prevalent in low-income rural populations, potentially excluding vulnerable groups from beneficial health interventions [99, 100]. Despite the clear advantages of mHealth/DH interventions, care must be taken during intervention development not to exclude populations and avoid exacerbating inequity and growing the ‘digital divide’ [101].

### Limitations

While this systematic review provides valuable evidence-based insights for the use of mHealth/DH interventions in SSA immunisation programmes, several key limitations must be acknowledged. Firstly, only published literature was included, highlighting potential publication bias. A more thorough search of the unpublished material or clinical trial registries would have reduced this limitation but was not possible due to resource constraints.

Additionally, it is recognised that in a narrative synthesis, although guided by quantitative findings, there is possibility for subjectivity in selection of findings which could have led to reporting bias, however, the certainty of evidence assessment considers the potential impact of reporting and publication bias on the confidence in presented results. In terms of study findings, it is recognised that by limiting the study context to only 19 SSA countries, the search breadth and inclusion criteria was limited and excluded countries such as South Africa, Ethiopia and Tanzania where mHealth/DH research is occurring [93]. Another limitation is generalisability of findings, with Nigeria being disproportionately represented here. Although Kenya, Burkina Faso and Cote D’Ivoire were represented, the overall finding’s transferability to these countries is also limited due to study low numbers. Although the included studies were from a mix of urban and rural settings, it must be noted that different countries and settings likely have different preferences and cultural factors for communications. For example, it was shown that Kenyan farmers preferred phone calls over SMS [83]. Finally, it is recognised that unique disease and context-specific factors must be considered, and caution made when transferring findings relating to DTP/Pentavalent vaccine to malaria vaccine context.

## Conclusion

Based on the findings of this review, a combined voice-based and SMS intervention is recommended to support the timely completion of the three-dose malaria vaccine schedule within SSA malaria immunisation programmes. While the available evidence highlights the potential of mHealth/DH interventions to improve vaccination uptake, these recommendations should be interpreted in the context of underlying certainty of evidence and the review’s outlined limitations. Nonetheless, the review’s evidence-based findings provide valuable insights for the design and implementation of future mHealth/DH interventions within immunisation programmes in SSA.

## Data Availability

All relevant data are within the manuscript and its Supporting Information files.

## Acknowledgements

The review team would like to express their deepest thanks to Marshall Dozier (Academic Support Librarian at University of Edinburgh’s College of Medicine and Veterinary Medicine) for aiding the review team in search strategy development. We would also like to thank Dr Eric Chen (Usher Institute) for his useful guidance throughout the systematic review process, and Dr Niall Anderson (Medical Statistician at Usher Institute, University of Edinburgh’s College of Medicine and Veterinary Medicine) for providing guidance and input regarding our quantitative findings and synthesis approach.

## Ethical Approval

No primary data was generated in this research project; therefore level 1 ethical approval was sought and granted by University of Edinburgh’s UMREG (Usher Masters’ Research Ethics Group).

## Supporting Information (See separate document)

**S1** PRISMA Checklist

**S2** PICOS, Inclusion/Exclusion Criteria

**S3** Search Strategies

**S4** Included Studies and Description of Intervention-Type Subgroups

**S5** GRADE Certainty of Evidence Assessment Explanations

**S6** Raw quantitative data for Forest Plots

