## Supplementary Information for "Systematic review investigating mHealth and digital health interventions for increasing vaccination uptake in 19 Sub-Saharan African countries: Recommendations for the malaria vaccine rollout"

**S1 – PRISMA Checklist**

| **Section and Topic** | **Item #** | **Checklist item** | **Location where item is reported** |
| --- | --- | --- | --- |
| **TITLE** | | |  |
| Title | 1 | Identify the report as a systematic review. | Page 1 |
| **ABSTRACT** | | |  |
| Abstract | 2 | See the PRISMA 2020 for Abstracts checklist. | Page 2 |
| **INTRODUCTION** | | |  |
| Rationale | 3 | Describe the rationale for the review in the context of existing knowledge. | Page 5/6 |
| Objectives | 4 | Provide an explicit statement of the objective(s) or question(s) the review addresses. | Page 6 |
| **METHODS** | | |  |
| Eligibility criteria | 5 | Specify the inclusion and exclusion criteria for the review and how studies were grouped for the syntheses. | Page 7 and S2 |
| Information sources | 6 | Specify all databases, registers, websites, organisations, reference lists and other sources searched or consulted to identify studies. Specify the date when each source was last searched or consulted. | Page 8 |
| Search strategy | 7 | Present the full search strategies for all databases, registers and websites, including any filters and limits used. | S3 |
| Selection process | 8 | Specify the methods used to decide whether a study met the inclusion criteria of the review, including how many reviewers screened each record and each report retrieved, whether they worked independently, and if applicable, details of automation tools used in the process. | Page 8 |
| Data collection process | 9 | Specify the methods used to collect data from reports, including how many reviewers collected data from each report, whether they worked independently, any processes for obtaining or confirming data from study investigators, and if applicable, details of automation tools used in the process. | Page 8 |
| Data items | 10a | List and define all outcomes for which data were sought. Specify whether all results that were compatible with each outcome domain in each study were sought (e.g. for all measures, time points, analyses), and if not, the methods used to decide which results to collect. | Page 7 |
|  | 10b | List and define all other variables for which data were sought (e.g. participant and intervention characteristics, funding sources). Describe any assumptions made about any missing or unclear information. | Table 1 and 2 |
| Study risk of bias assessment | 11 | Specify the methods used to assess risk of bias in the included studies, including details of the tool(s) used, how many reviewers assessed each study and whether they worked independently, and if applicable, details of automation tools used in the process. | Page 8 |
| Effect measures | 12 | Specify for each outcome the effect measure(s) (e.g. risk ratio, mean difference) used in the synthesis or presentation of results. | Page 10 |
| Synthesis methods | 13a | Describe the processes used to decide which studies were eligible for each synthesis (e.g. tabulating the study intervention characteristics and comparing against the planned groups for each synthesis (item #5)). | Page 9 and S2 |
|  | 13b | Describe any methods required to prepare the data for presentation or synthesis, such as handling of missing summary statistics, or data conversions. | Page 9 |
|  | 13c | Describe any methods used to tabulate or visually display results of individual studies and syntheses. | Page 9 |
|  | 13d | Describe any methods used to synthesize results and provide a rationale for the choice(s). If meta-analysis was performed, describe the model(s), method(s) to identify the presence and extent of statistical heterogeneity, and software package(s) used. | Page 8/9 |
|  | 13e | Describe any methods used to explore possible causes of heterogeneity among study results (e.g. subgroup analysis, meta-regression). | Page 9 |
|  | 13f | Describe any sensitivity analyses conducted to assess robustness of the synthesized results. | Page 9 |
| Reporting bias assessment | 14 | Describe any methods used to assess risk of bias due to missing results in a synthesis (arising from reporting biases). | Page 8/9 |
| Certainty assessment | 15 | Describe any methods used to assess certainty (or confidence) in the body of evidence for an outcome. | Page 8/9 |
| **RESULTS** | | |  |
| Study selection | 16a | Describe the results of the search and selection process, from the number of records identified in the search to the number of studies included in the review, ideally using a flow diagram. | Page 10 and Fig 1 |
|  | 16b | Cite studies that might appear to meet the inclusion criteria, but which were excluded, explain why they were excluded. | Corresponding author |
| Study characteristics | 17 | Cite each included study and present its characteristics. | Table 1 and S4 |
| Risk of bias in studies | 18 | Present assessments of risk of bias for each included study. | Page 21 and Fig 3 |
| Results of individual studies | 19 | For all outcomes, present, for each study: (a) summary statistics for each group (where appropriate) and (b) an effect estimate and its precision (e.g. confidence/credible interval), ideally using structured tables or plots. | Table 2 |
| Results of syntheses | 20a | For each synthesis, briefly summarise the characteristics and risk of bias among contributing studies. | Pages 21 to 31 |
|  | 20b | Present results of all statistical syntheses conducted. If meta-analysis was done, present for each the summary estimate and its precision (e.g. confidence/credible interval) and measures of statistical heterogeneity. If comparing groups, describe the direction of the effect. | Pages 26 to 31 |
|  | 20c | Present results of all investigations of possible causes of heterogeneity among study results. | Pages 21 to 23 |
|  | 20d | Present results of all sensitivity analyses conducted to assess the robustness of the synthesized results. | Pages 21 to 31 |
| Reporting biases | 21 | Present assessments of risk of bias due to missing results (arising from reporting biases) for each synthesis assessed. | 21 and Fig 3 |
| Certainty of evidence | 22 | Present assessments of certainty (or confidence) in the body of evidence for each outcome assessed. | Page 23 and Table 3 |
| **DISCUSSION** | | |  |
| Discussion | 23a | Provide a general interpretation of the results in the context of other evidence. | Pages 32 to 38 |
|  | 23b | Discuss any limitations of the evidence included in the review. | Page 40 |
|  | 23c | Discuss any limitations of the review processes used. | Page 40 |
|  | 23d | Discuss implications of the results for practice, policy, and future research. | Page 32 to 40 |
| **OTHER INFORMATION** | | |  |
| Registration and protocol | 24a | Provide registration information for the review, including register name and registration number, or state that the review was not registered. | Page 7 |
|  | 24b | Indicate where the review protocol can be accessed, or state that a protocol was not prepared. | Page 7 |
|  | 24c | Describe and explain any amendments to information provided at registration or in the protocol. | Prospero |
| Support | 25 | Describe sources of financial or non-financial support for the review, and the role of the funders or sponsors in the review. | Page 40 |
| Competing interests | 26 | Declare any competing interests of review authors. | N/A |
| Availability of data, code and other materials | 27 | Report which of the following are publicly available and where they can be found: template data collection forms; data extracted from included studies; data used for all analyses; analytic code; any other materials used in the review. | Supplementary info or corresponding author |

**S2** – **PICOSS Framework and Inclusion/Exclusion Criteria**

**S2.1** The PICOSS framework used to guide this systematic review.

| **Aspect of PICOSS** | **The PICOSS used** |
| --- | --- |
| Population | Mothers/caregivers of children or infants (<5 years old) that are receiving the DTP or Pentavalent vaccines in the 19 SSA countries or HCWs that are involved in administering the DTP or Pentavalent vaccines. |
| Intervention | mHealth/DH interventions that are used in essential childhood immunisation programmes in SSA to increase vaccination uptake or coverage |
| Comparison | Non-digital strategies for increasing vaccination uptake in SSA (e.g. Physical or verbal appointment reminders). |
| Outcome | Vaccination Uptake or Coverage Rate (%), Vaccination Schedule Completion (%), Vaccination Schedule Timeliness Rate (%), Risk Ratio (RR) or Odds Ratio (OR) |
| Setting | 19 Countries in SSA (Benin, Burkina Faso, Burundi, Cameroon, Central African Republic, Chad, Cote d’Ivoire, Democratic Republic of Congo, Ghana, Guinea, Kenya, Liberia, Malawi, Mozambique, Niger, Nigeria, Sierra Leone, South Sudan and Uganda) |
| Study Design | Experimental Studies (Randomised Control Trials [RCTs] or Non-Randomised Control Trials [Non-RCTs]) or Pre-post case studies (Technology implementation or assessment study) |

**S2.2** PICOSS guided inclusion/exclusion criteria for assessing study inclusion eligibility.

| **PICOSS** | **Inclusion** | **Exclusion** |
| --- | --- | --- |
| Population | Any study with a study population receiving or administering the DTP or Pentavalent Vaccine. | Any studies that report results from other immunisation programmes for example, COVID-19 vaccines or Human Papillomavirus (HPV) |
| Intervention | Studies which implement an mHealth or Digital Health intervention to increase vaccination uptake. For example, automated-mobile phone appointment reminders. | Studies which implement non-digital interventions for increasing vaccination uptake. For example, physical or verbal appointment reminders. |
| Comparison | Non-digital interventions for increasing vaccine uptake (comparator arm for experimental studies) | Non-digital interventions are used to increase vaccine uptake in the immunisation programmes. |
| Outcome | Studies reporting vaccination uptake rate (%), schedule completion (%), vaccination timeliness rate (%), RR or OR. | N/A |
| Setting | 19 Countries in SSA rolling out malaria vaccine in 2024. | Other SSA countries that will not be rolling out malaria vaccine eg. South Africa |
| Study Design | Only epidemiological experimental studies (RCTs or Non-RCTs) or Technology implementation or assessment case studies. | Observational studies such as case-control or cross-sectional studies, other types of studies including SRs and RCT protocols. |

**S3 - Search Strategies**

**S3.1** The key search terms and four concept area clusters used to make up the search

| **Concept Area Clusters** | **Search Terms** |
| --- | --- |
| mHealth and Digital Health Interventions | digital health OR mhealth OR m-health OR ehealth OR e-health OR Cell Phone OR Smartphone OR mobile device OR laptop OR computer OR electronic OR technology OR telemedicine OR Communication technology OR Text Messaging OR Text* OR Short Message OR SMS OR mobile app* OR application OR software OR App* OR App-based OR Web OR Web-based OR Internet* OR Digital* OR WhatsApp OR social media |
| Childhood Immunisation Programmes | Vaccination* OR immunization* OR immunisation* OR vaccine* OR DTP vaccine OR Diphtheria-Pertussis-Tetanus vaccine OR Pentavalent vaccine OR Reminder System OR Appointment OR Scheduling |
| Outcomes (Vaccination Uptake) and Study Design | Uptake OR coverage OR completion OR Timeliness OR Trial OR Implementation OR intervention OR reminder OR appointment |
| Setting | Benin OR Burkina Faso OR Burundi OR Cameroon OR Central African Republic OR Chad OR Cote d’Ivoire OR Democratic Republic of Congo OR Ghana OR Guinea OR Kenya OR Liberia OR Malawi OR Mozambique OR Niger OR Nigeria OR Sierra Leone OR South Sudan OR Uganda |

**Grey Literature**

Grey Literature search comprised search sources OpenHIA (OpenHIA.org), OpenMRS (OpenMRS.org) and WHO’s mHealth/DH working group publications (www.who.int/teams/digital-health-and-innovation). Only basic single keyword searches were conducted on these websites and were only related to immunisations cluster (‘immunisation’ or ‘immunization’ or ‘vaccine’ or ‘vaccination’).

**Africa Journals Online (AJOL)**

vaccination* OR immunization* OR immunisation* OR vaccine*

AND digital health OR mhealth OR m-health OR ehealth OR e-health OR Cell Phone OR smartphone OR mobile device OR telemedicine OR communication OR Text Messaging OR Short Message OR sms OR mobile app* OR app* OR app-based OR web OR web-based OR internet* OR digital* OR whatsapp OR social media

**Africa Index Medicus (AIM)**

digital health OR mhealth OR ehealth OR Cell Phone OR Smartphone OR mobile device OR telemedicine OR Communication OR Text Messaging OR Short Message OR SMS OR mobile app* OR application OR App* OR App-based OR Web OR Web-based OR Internet* OR WhatsApp OR social media

AND Cluster Vaccination* OR immunization* OR immunisation* OR vaccine* OR Reminder System* OR Appointment Schedul* OR DTP Vaccin* OR Diphtheria-Pertussis-Tetanus vaccin*

**Cochrane Central**

#1	(digital health):ti,ab,kw

#2	(mHealth):ti,ab,kw

#3	(eHealth):ti,ab,kw

#4	(cell phone):ti,ab,kw

#5	(smart phone):ti,ab,kw

#6	(mobile device):ti,ab,kw

#7	(laptop):ti,ab,kw

#8	(computer):ti,ab,kw

#9	(electronic):ti,ab,kw

#10	(technology):ti,ab,kw

#11	(telemedicine):ti,ab,kw

#12	(Communication):ti,ab,kw

#13	(text messaging):ti,ab,kw

#14	(short message):ti,ab,kw

#15	(SMS):ti,ab,kw

#16	(Mobile App*):ti,ab,kw

#17	(application):ti,ab,kw

#18	(software):ti,ab,kw

#19	(App based):ti,ab,kw

#20	("web"):ti,ab,kw

#21	(Web based):ti,ab,kw

#22	(internet):ti,ab,kw

#23	(social media):ti,ab,kw

#24	(digital):ti,ab,kw

#25	(WhatsApp):ti,ab,kw

#26	(Text*):ti,ab,kw

#27	(App*):ti,ab,kw

#28	{OR #1-#27}

#29	(Vaccination*):ti,ab,kw

#30	(Immunization*):ti,ab,kw

#31	(Immunisation*):ti,ab,kw

#32	(Vaccine*):ti,ab,kw

#33	(DTP vaccin*):ti,ab,kw

#34	(Diphtheria-Pertussis-Tetanus vaccin*):ti,ab,kw

#35	(Pentavalent Vaccin*):ti,ab,kw

#36	Reminder System*

#37	Appointment Schedul*

#38	{OR #29-#37}

#39	#28 AND #38

#40	(Uptake OR Coverage OR Completion OR Timeliness OR Trial OR Implementation OR Intervention):ti,ab,kw

#41	(Benin OR Burkina Faso OR Burundi OR Cameroon OR Central African Republic OR Chad OR Cote d’Ivoire OR Democratic Republic of Congo OR Ghana OR Guinea OR Kenya OR Liberia OR Malawi OR Mozambique OR Niger OR Nigeria OR Sierra Leone OR South Sudan OR Uganda):ti,ab,kw

#42	#39 AND #40 AND #41

**Embase (Ovid)**

1.	electronic medical record/ or medical information/ or digital health/ or Internet/

2.	mobile phone/ or mobile application/ or telemedicine/ or mhealth.mp. or mobile health application/

3.	smartphone/

4.	microcomputer/ or laptop/

5.	ehealth.mp. or telehealth/

6.	medical electronics/

7.	digital health technology/ or digital technology/

8.	communication technology/ or telecommunication/

9.	automation/ or text messaging/

10.	social media/

11.	WhatsApp.mp.

12.	WhatsApp.mp.

13.	1 or 2 or 3 or 4 or 5 or 6 or 7 or 8 or 9 or 10 or 11 or 12

14.	vaccination/ or vaccination coverage/

15.	immunization/ or mass immunization/

16.	Immunisation.mp.

17.	vaccine hesitancy/ or vaccine/

18.	DTP vaccine.mp. or diphtheria pertussis tetanus vaccine/

19.	Pentavalent vaccin*.mp.

20.	diphtheria pertussis tetanus Haemophilus influenzae type b hepatitis B vaccine/

21.	reminder system/

22.	appointment schedul*.mp. [mp=title, abstract, heading word, drug trade name, original title, device manufacturer, drug manufacturer, device trade name, keyword heading word, floating subheading word, candidate term word]

23.	14 or 15 or 16 or 17 or 18 or 19 or 20 or 21 or 22

24.	(Benin or Burkina Faso or Burundi or Cameroon or Central African Republic or Chad or Cote d'Ivoire or Democratic Republic of Congo or Ghana or Guinea or Kenya or Liberia or Malawi or Mozambique or Niger or Nigeria or Sierra Leone or South Sudan or Uganda).mp. [mp=title, abstract, heading word, drug trade name, original title, device manufacturer, drug manufacturer, device trade name, keyword heading word, floating subheading word, candidate term word]

25.	13 and 23 and 24

**Global Health (Ovid)**

1.	digital health.mp. or digital technology.sh. or social media.sh.

2.	(technology or mobile equipment).sh. or mHealth/ or communication.sh.

3.	ehealth.mp. or e-health/

4.	smartphone.mp. or mobile telephones/

5.	cell phone.mp.

6.	mobile applications.sh.

7.	computers/

8.	electronics/

9.	telemedicine/ or telecommunications.sh.

10.	text messaging/

11.	internet/

12.	social media/

13.	WhatsApp.mp.

14.	1 or 2 or 3 or 4 or 5 or 6 or 7 or 8 or 9 or 10 or 11 or 12 or 13

15.	vaccination/ or mass vaccination/

16.	immunization/ or immunization programmes/

17.	immunisation.mp.

18.	(vaccines or diphtheria pertussis tetanus vaccines).sh.

19.	pentavalent vaccin*.mp.

20.	reminder system.mp.

21.	appointment schedul*.mp. [mp=abstract, title, original title, broad terms, heading words, cabicodes words]

22.	15 or 16 or 17 or 18 or 19 or 20 or 21

23.	(Benin or Burkina Faso or Burundi or Cameroon or Central African Republic or Chad or Cote d'Ivoire or Democratic Republic of Congo or Ghana or Guinea or Kenya or Liberia or Malawi or Mozambique or Niger or Nigeria or Sierra Leone or South Sudan or Uganda).mp. [mp=abstract, title, original title, broad terms, heading words, cabicodes words]

24.	14 and 22 and 23

**Medline (Ovid)**

1.	Digital Health/ or Internet/ or Mobile Applications/ or Telemedicine/

2.	mHealth.mp.

3.	ehealth.mp.

4.	Cell Phone/ or Smartphone/

5.	microcomputers/ or computers, handheld/ or minicomputers/

6.	electronics/ or digital technology/ or electronics, medical/

7.	communication/ or "cell phone use"/

8.	Text Messaging/

9.	Social Media/

10.	WhatsApp.mp.

11.	1 or 2 or 3 or 4 or 5 or 6 or 7 or 8 or 9 or 10

12.	Vaccination/ or Mass Vaccination/ or Vaccination Hesitancy/ or Vaccination Coverage/

13.	Immunization Programs/ or Immunization/ or Immunization Schedule/

14.	Immunisation.mp.

15.	Vaccines/

16.	Diphtheria-Tetanus-Pertussis Vaccine/

17.	Pentavalent vaccine.mp.

18.	"Appointments and Schedules"/ or Reminder Systems/

19.	12 or 13 or 14 or 15 or 16 or 17 or 18

20.	(Benin or Burkina Faso or Burundi or Cameroon or Central African Republic or Chad or Cote d'Ivoire or Democratic Republic of Congo or Ghana or Guinea or Kenya or Liberia or Malawi or Mozambique or Niger or Nigeria or Sierra Leone or South Sudan or Uganda).mp. [mp=title, book title, abstract, original title, name of substance word, subject heading word, floating sub-heading word, keyword heading word, organism supplementary concept word, protocol supplementary concept word, rare disease supplementary concept word, unique identifier, synonyms, population supplementary concept word, anatomy supplementary concept word]

21.	11 and 19 and 20

**Scopus**

(TITLE-ABS-KEY(benin OR {burkina faso} OR burundi OR cameroon OR {central african republic} OR chad OR {cote d'ivoire} OR {democratic republic of congo} OR ghana OR guinea OR kenya OR liberia OR malawi OR mozambique OR niger OR nigeria OR {sierra leone} OR {south sudan} OR uganda))

AND (TITLE-ABS-KEY(uptake OR coverage OR completion OR timeliness OR trial OR implementation OR intervention))

AND (TITLE-ABS-KEY(Vaccination* OR immunization* OR immunisation* OR vaccine* OR {DTP vaccine} OR {Diphtheria-Pertussis-Tetanus vaccine} OR {Pentavalent vaccine} OR {Reminder System} OR "Appointment Schedul*" ))

AND (TITLE-ABS-KEY( {digital health} OR mhealth OR m-health OR ehealth OR e-health OR {Cell Phone} OR smartphone OR {mobile device} OR laptop OR computer OR electronic OR technology OR telemedicine OR communication OR {Text Messaging} OR text* OR {Short Message} OR sms OR "mobile app*" OR application OR software OR app* OR app-based OR message OR web OR web-based OR internet OR whatsapp OR {social media} ))

**S4 – Included Studies and Description of Intervention-Type Subgroups**

| **Author and Year** | **Location** | **Title** |
| --- | --- | --- |
| Brown and Oluwatosin, 2017 [52] | Nigeria | Feasibility of implementing a cellphone-based reminder/recall strategy to improve childhood routine immunization in a low-resource setting: a descriptive report |
| Brown *et al*, 2016 [53] | Nigeria | Effects of Community Health Nurse-Led Intervention on Childhood Routine Immunization Completion in Primary Health Care Centers in Ibadan, Nigeria |
| Dissieka *et al*, 2019 [54] | Cote D'Ivoire | Providing mothers with mobile phone message reminders increases childhood immunization and vitamin A supplementation coverage in Cote d'Ivoire: a randomized controlled trial. |
| Ekhaguere *et al*, 2019 [55] | Nigeria | Automated phone call and text reminders for childhood immunisations (PRIMM): a randomised controlled trial in Nigeria |
| Eze and Adeleye, 2015 [56] | Nigeria | Enhancing routine immunization performance using innovative technology in an urban area of Nigeria |
| Gibson *et al*, 2017 [57] | Kenya | Mobile phone-delivered reminders and incentives to improve childhood immunisation coverage and timeliness in Kenya (M-SIMU): a cluster randomised controlled trial. |
| Haji *et al*, 2016 [58] | Kenya | Reducing routine vaccination dropout rates: evaluating two interventions in three Kenyan districts, 2014 |
| Ibraheem *et al*, 2021 [59] | Nigeria | Effects of call reminders, short message services (SMS) reminders, and SMS immunization facts on childhood routine vaccination timing and completion in Ilorin, Nigeria. |
| Kawakatsu *et al*, 2020 [60] | Nigeria | Cost-effectiveness of SMS appointment reminders in increasing vaccination uptake in Lagos, Nigeria: A multi-centered randomized controlled trial. |
| Oladepo *et al*, 2020 [61] | Nigeria | Outcome of reminder text messages intervention on completion of routine immunization in rural areas, Nigeria. |
| Sampson *et al*, 2023 [62] | Nigeria | An assessment of the effectiveness of an electronic wristband in improving routine immunization timeliness and reducing drop-out |
| Schlumberger *et al*, 2015 [63] | Burkina Faso | Positive impact on the Expanded Program on Immunization when sending call-back SMS through a Computerized Immunization Register, Bobo Dioulasso (Burkina Faso) |
| Yunusa *et al*, 2022 [64] | Nigeria | Effect of mobile phone text message and call reminders in the completeness of pentavalent vaccines in Kano state, Nigeria. |
| Yunusa *et al*, 2024 [65] | Nigeria | Utilization of Mobile Reminders in Improving the Completeness and Timeliness of Routine Childhood Immunization in Kano Metropolis, Nigeria: A Randomized Controlled Trial |

**S4.1** The author and year, study location and title of the 14 included studies.

**S4.2** Description of mHealth/DH intervention-types subgroup and which studies are included in each.

| **mHealth/DH Intervention-type Subgroup Investigated** | **Definition** | **Which Included Studies Are Related to Each Category** |
| --- | --- | --- |
| SMS-Only Appointment Reminders | Interventions that only comprised SMS appointment reminders. For example, Ibraheem *et al* (2021) sent this SMS to participants: “Dear parent, your child is due for the next vaccines tomorrow. Please bring your child for vaccination at the hospital at 8 am. Thank you.” [59]. | Seven studies included SMS-only interventions: Eze and Adeleye (2015) [56], Gibson *et al* (2017) [57], Haji et al (2016) [58], Ibraheem et al (2021) [59], Kawakatsu et al (2020) [60], Schlumberger *et al* (2015) [63], Yunusa *et al* (2022) [64]. |
| ‘SMS-Plus’ Interventions | These are SMS appointment reminder messages that have been ‘enhanced’. Either in combination with small cash incentives or provided additional info such as educational messages about immunisations. | Three studies were classed as ‘SMS-Plus’ interventions: Gibson *et al* (2017) [57], Ibraheem *et al* (2021) [59], Oladepo *et al* (2020) [61]. Gibson *et al* (2017) combined SMS with 75KES (Group B) and 200KES (Group C). Participants in Oladepo *et al* (2020) and Group C in Ibraheem *et al* (2021), received educational SMS messages. |
| ‘SMS and/or Voice Messages or Phone Calls’ | This subgroup refers to SMS appointment reminders either in conjunction with a voice-based component (voice message or phone call), or participants were given the choice between receiving SMS or voice message (allowing recipients to hear reminder). | Three studies were included in this subgroup: Dissieka *et al* (2019) [54], Ekhaguere *et al* (2019) [55], Yunusa *et al* (2024) [65]. Dissieka *et al* (2019) offered the choice of receiving SMS or voice message reminders. Ekhaguere *et al* (2019) combined SMS and voice messages, and Yunusa *et al* (2024) combined SMS with phone calls. |
| ‘Phone Call-Only reminders’ | These involved mothers/caregivers being called to remind them about their child’s upcoming immunisations (solely voice-based). | Three studies were included in this subgroup: Brown and Oluwatosin (2017) [52], Brown *et al* (2016) [53], Ibraheem *et al* (2021) [59]. Brown and Oluwatosin (2017), Group A in Brown *et al* (2016) and Group A in Ibraheem *et al* (2021) solely investigated phone call reminders. Group C in Brown *et al* (2016) investigated phone call reminders combined with specialist vaccinator/HCW training (not mHealth) |
| ‘Wearable Electronic Immunisation Alert Wristband’ | This was a wearable electronic wristband worn by mother/caregivers; it was digitally programmed to flash in the lead up to the day of a child’s immunisation appointment. The flashing red light could be easily spotted in the mother/caregivers’ hands. It flashed five times throughout the intervention, at week 0, week 6 (Penta1), week 10 (Penta2), week 14 (Penta3) and 9months. | Only Sampson *et al* (2023) [62]. |

**S5 – GRADE Certainty of Evidence Assessment Explanations**

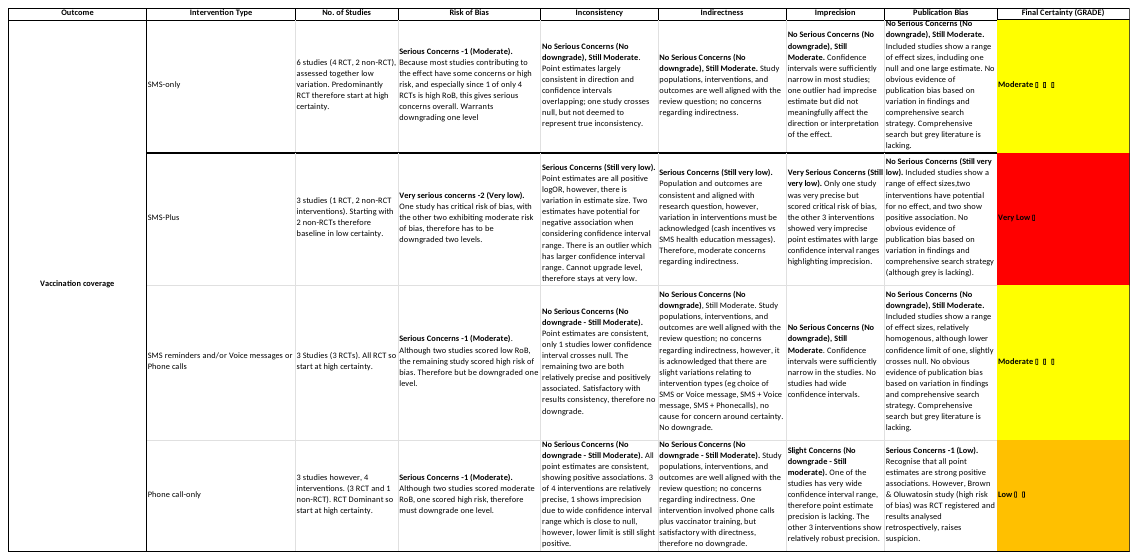
**S5.3** GRADE Certainty of evidence assessment for vaccination coverage

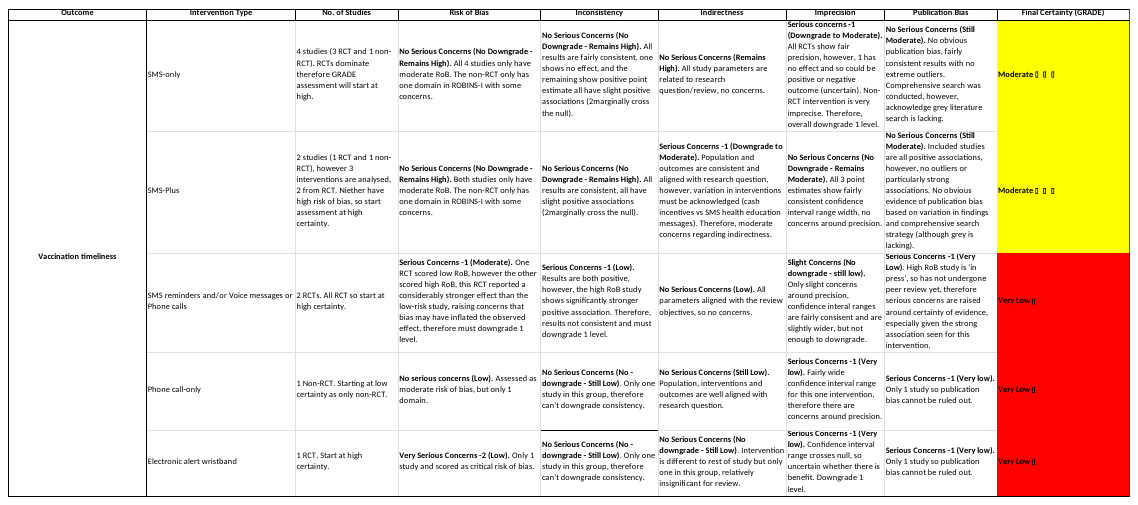
**S5.4** GRADE Certainty of evidence assessment for vaccination timeliness

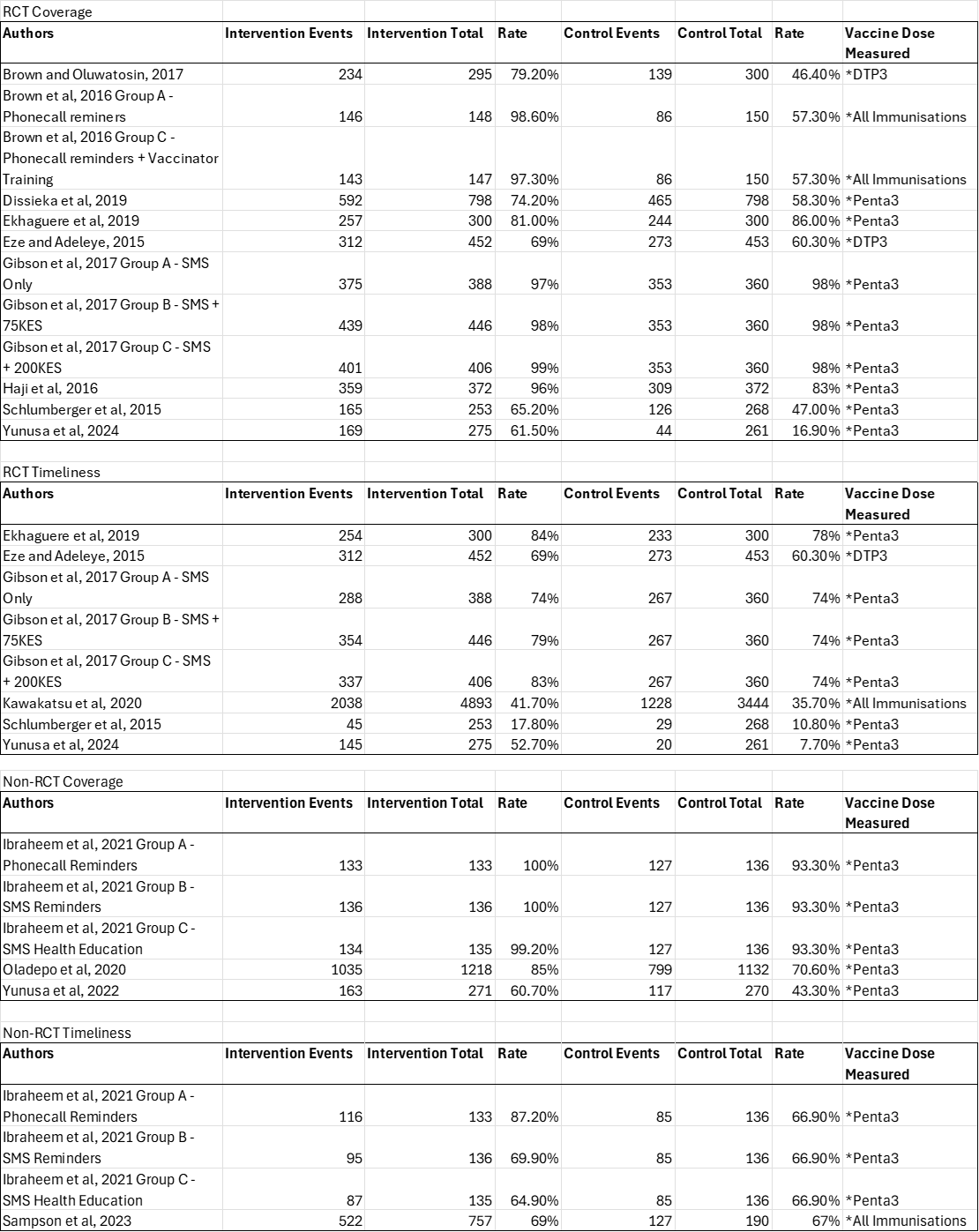
**S6 – Raw Quantitative Data used to calculate Forest Plots and LogOR**
